# EMOD with Full Parasite Genetics: A modeling framework for evaluating parasite genetic metrics for operational malaria molecular surveillance

**DOI:** 10.64898/2026.06.05.26355027

**Authors:** Jessica V. Ribado, Joshua Suresh, Daniel Bridenbecker, Jonathan R. Russell, Albert Lee, Edward Wenger, Guillaume Chabot-Couture, Joshua L. Proctor, Katherine E. Battle, Caitlin A. Bever

## Abstract

Malaria molecular surveillance (MMS) is becoming increasingly common in endemic settings and has been proposed as a tool for monitoring parasite transmission to inform programmatic decision-making. However, the conditions under which parasite genetic metrics provide interpretable signals for broader use cases, such as assessing intervention impacts and detecting importation, remain under-characterized. We present EMOD with Full Parasite Genetics (FPG), a simulation framework designed to explore how parasite genetic metrics arise from transmission, intervention, importation, and sampling processes at programmatically relevant timescales. Using seasonal scenarios across a range of transmission intensities, we demonstrate three principal findings. First, genetic metrics can detect insecticide-treated net intervention impacts at seasonal and yearly timescales, but the strength, timing, and form of the relationship between genetic and epidemiological measures vary by metric and sampling timing. Second, importation can break the expected relationship between parasite genetic diversity from local transmission intensity at very low incidence, allowing low-transmission settings with substantial importation to maintain elevated diversity metrics. Third, convenience sampling practices, including sample size, collection timing, and the clinical composition of sampled populations, introduce non-random biases in genetic metric estimation in a way that obscures the true transmission signal. Together, these findings show that parasite genetic metrics can support operational surveillance, but that their interpretation depends on transmission context, importation, metric choice, and sampling design. EMOD FPG provides a framework for evaluating these dependencies in future setting-specific analyses and for guiding the interpretation of parasite genetic data across sites and over time.

**Author summary:** Malaria control programs are increasingly interested in using parasite genetic data to understand where transmission is changing, whether interventions are working, and whether infections are being imported from elsewhere. However, genetic data can be difficult to interpret because the patterns observed in sampled parasites are shaped not only by transmission, but also by importation, seasonality, and how samples are collected.

We developed a simulation model that tracks malaria parasite genomes through human infections, mosquito transmission, recombination, and field-like sampling. We used this model to evaluate how parasite genetic metrics behave under realistic programmatic conditions. We found that genetic metrics can detect the effects of an insecticide-treated net intervention campaign, but that the timing and strength of the signal depend on the metric used. We also found that importation can make low-transmission settings appear genetically diverse, complicating interpretation of genetic metrics as sole indicators of transmission. Finally, we showed that sample size, collection timing, and whether samples come from symptomatic or asymptomatic infections can systematically bias genetic estimates.

These findings suggest that parasite genetic data can support malaria surveillance, but only when interpreted through careful consideration of transmission context, importation, metric choice, and sampling design.

## Introduction

Malaria molecular surveillance (MMS) has increased substantially over the past decade with improved capacity, evolving from proof-of-concept work in research settings toward programmatic integration with national malaria control programs (NMCPs) [1,2]. The most established programmatic applications of MMS include monitoring molecular markers of antimalarial drug resistance and detecting deletions in *pfhrp2/3* that can cause false negative *Plasmodium falciparum* rapid diagnostic tests (RDTs)[3–5]. Beyond these established applications, there is growing interest in using parasite genetic data to support broader epidemiological questions, as highlighted in a 2019 WHO Technical Consultation on the role of parasite and vector genetic data in malaria surveillance [6]. These use cases include measuring changes in transmission, evaluating intervention impact, and identifying parasite importation.

Empirical studies have demonstrated that parasite genetic diversity metrics, such as the proportion of polygenomic infections, complexity of infection (COI), and identity-by-descent (IBD) among parasite strains may correlate with measures of transmission intensity like clinical incidence or parasite prevalence. In Senegal, the proportion of polygenomic infections was identified as a strong genetic correlate of reported malaria incidence across diverse transmission settings [7,8]. Likewise, in Uganda, measures of genetic diversity from high-resolution microhaplotype data reflected spatial variation in prevalence data more strongly than clinical incidence, which is shaped by population immunity, treatment seeking, and other factors [9]. Genetic metrics have also been used to characterize connectivity and quantify importation rates across multiple African settings [10–12]. Studies like these demonstrate the promise of MMS for surveillance questions beyond resistance monitoring.

However, these same findings raise important questions about interpretation. The relationship between parasite genetic metrics and epidemiological transmission measures is not fixed across settings. Transmission intensity, seasonality, importation, intervention history, sampling timing, sample size, and the clinical composition of sampled infections may all influence the genetic metrics observed. As a result, the same genetic metric cannot be interpreted the same way across settings without accounting for these various contextual factors. This creates a central challenge for programmatic MMS: determining when genetic metrics provide reliable information for specific surveillance use cases, and when they may be misleading without additional context.

Mathematical models provide a way to examine these relationships under controlled conditions. Existing malaria genetic epidemiology models have clarified how transmission intensity and population genetic processes shape parasite diversity [13–15]. However, many models simplify the intervention dynamics, within-host biology, spatial structure, and sampling processes that are central to programmatic MMS use cases. These simplifications are particularly relevant when asking whether genetic metrics reflect intervention impact, local versus imported transmission, or clinical versus asymptomatic sampling, because each of these questions depends on processes such as realistic intervention effects, symptom-generating infection dynamics, cotransmission and superinfection, spatial connectivity, and sample selection for sequencing.

Building on existing modeling frameworks (see Supplemental Table 1, [13–15]), EMOD with Full Parasite Genetics (FPG) was developed to address these questions through a framework that mechanistically links parasite genetic surveillance observations to the epidemiological processes that generate them. FPG layers parasite genetics onto EMOD, a highly detailed agent-based malaria transmission model that has been applied extensively to evaluate intervention strategies, estimate transmission intensity, and project elimination scenarios across diverse epidemiological settings [16–21]. FPG explicitly tracks parasite genomes across human and mosquito infections, allowing genetically distinct strains to be transmitted, maintained within hosts, sampled by mosquitoes, recombined during oocyst formation, and transmitted onward. The framework also supports observational sampling schemes designed to mimic field parasite genetic data collection. This allows genetic metrics to be evaluated not only as properties of the simulated parasite population, but also as quantities estimated through realistic sampling processes.

We use FPG to evaluate three interconnected questions relevant to MMS. First, we test whether parasite genetic metrics can detect the impact of insecticide-treated net distribution in a seasonal transmission setting. Second, we examine how importation alters the relationship between genetic metrics and local transmission intensity, particularly at low incidence. Third, we quantify how sampling design, specifically sample size, collection timing, and clinical composition of sampled infections, affects genetic metric estimation. Together, these analyses show that parasite genetic metrics can support operational surveillance, but that their interpretation depends on intervention history, importation context, metric choice, and sampling design. FPG provides a framework for evaluating these dependencies in future setting-specific analyses.

## Methods

### Model overview

EMOD with Full Parasite Genetics (FPG) extends EMOD, a stochastic agent-based model of malaria transmission [16,22], by explicitly tracking parasite genomes through human infections, mosquito infections, and onward transmission (Figure 1A). In contrast to EMOD’s aggregate contagion-pool representation, FPG records individual human-vector transmission events, allowing transmission histories and parasite lineages to be reconstructed explicitly.

**Figure 1.**
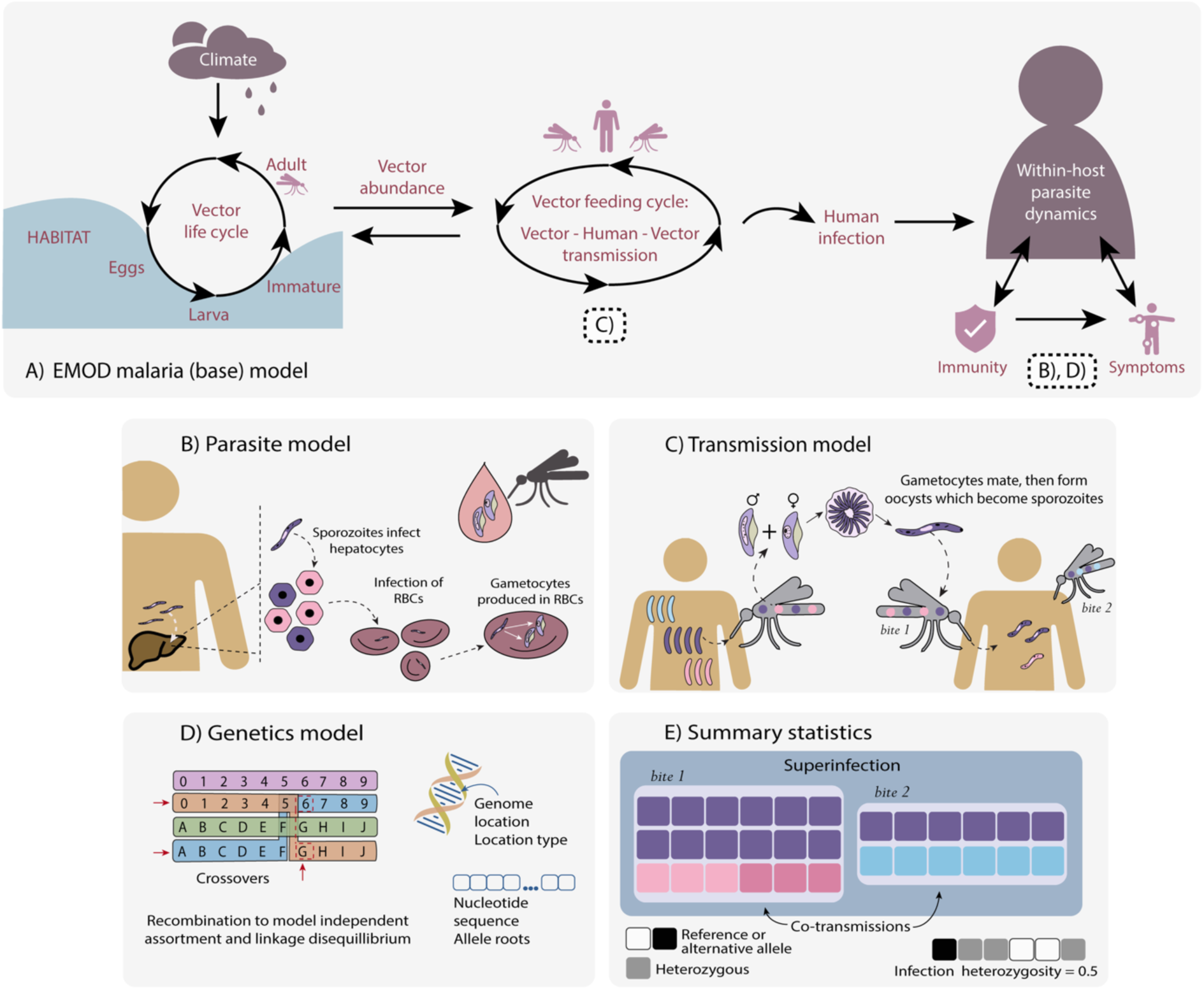
Overview of the EMOD malaria model and simulation framework for generating and characterizing within-host parasite genetic diversity. **(A) Epidemiological architecture** EMOD is an agent-based model in which each node contains populations of individual humans and vectors. Each simulated individual maintains their own infections, susceptibility, and interventions state. Infections are exchanged between humans and vectors, and interventions can be applied to both host populations. **(B) Parasite maturation** The parasite lifecycle is modeled across three compartments: infection (human host), transmission, and vector. Sporozoites initiate infection within the human by establishing hepatocytes, which then develop into infected red blood cells (iRBCs). iRBCs express surface antigens that drive antibody responses, with antigenic diversity and immunity governing the dynamics of new and existing infections. Mature gametocytes are produced and sampled by mosquito vectors during a blood meal. Within the vector, ingested gametocytes mate and undergo meiotic recombination in the oocyst stage before developing into sporozoites that are transmitted back to humans. **(C) Transmission** At each timestep, individual vectors select humans to bite based on intervention coverage and individual biting risk. Vectors sample male and female gametocytes from the bitten person’s circulating gametocyte pool, with genomic composition drawn in proportion to strain-specific gametocyte densities. Sampled gametocytes mate and progress through oocyst and sporozoite stages within the vector. Sporozoites are selected for transmission in proportion to the abundance of each sporozoite genotype cohort. **(D) Parasite genomes and recombination** Parasite genomes are modeled on the 14-chromosome structure of *P. falciparum*. Users may choose to incorporate known marker loci including *var* genes, *kelch13*, *hrp2*, *msp1*, *hrp3*, *crt*, *mdr1*, *dhfr*, *msp2*, and SNP identifiers. The model genome stores variant location, locus type (var genes or neutral alleles), ancestry (“roots”), and assigned alleles (“genotypes”) as variable-length arrays, allowing users to specify which attributes and loci to track. Recombination is implemented as a two-step process: an initial obligate crossover is placed with uniform probability along each chromosome pair, and secondary crossovers are drawn from a gamma distribution. Chromosome pairs for crossing over are selected uniformly, and recombinant chromosomes are combined to form new offspring genomes. This framework realistically captures both independent assortment and linkage disequilibrium across loci. **(E) Genetic metric calculations from simulated genomes** Four summary statistics are computed from the simulated parasite genomes. Ancestry root and genotype representations can be used to identify strains at different levels of realism. Using both genome representations, the model provides an upper bound of strain representations, while using genotypes only provides a more realistic lower bound of strains that may be detected from sequencing. Illustrative genetic metrics are shown, but for full genetic metric options and detail calculations see Table 1.

FPG tracks genetically distinct parasite strains within hosts and vectors, enabling explicit representation of cotransmission, superinfection, clonal propagation, and mosquito-stage recombination. Parasite genomes are linked to EMOD’s existing within-host infection dynamics, including parasite densities, gametocyte production, clinical symptoms, and acquired immunity. In mosquitoes, ingested gametocytes can form oocysts, undergo recombination when genetically distinct parental genomes are present, and generate sporozoite cohorts that may be transmitted onward.

The framework also includes an observational sampling model that generates simulated parasite genetic datasets under field-like sampling schemes. Together, these extensions allow FPG to evaluate how parasite genetic metrics are shaped by transmission intensity, intervention effects, importation, within-host diversity, recombination, and sampling design.

### Parasite genome representation

FPG represents each *P. falciparum* parasite genome using user-specified loci distributed across the parasite’s 14 nuclear chromosomes. At each tracked locus, the model stores both an allele value and an allele root. The allele value represents the observed genotype at that position, while the allele root records the ancestral infection from which that allele descends. This parallel representation allows the model to distinguish identity-by-state, based on shared allele values, from simulated identity-by-descent, based on shared ancestry.

The framework can represent multiple classes of loci, including neutral markers, immune-relevant loci, drug-resistance markers, and *pfhrp2/pfhrp3* deletions. In the present study, we focus on a 100-locus biallelic neutral SNP barcode, with loci distributed across chromosomes approximately in proportion to chromosome length. These loci do not affect parasite fitness, within-host dynamics, or immune responses in the simulations analyzed here.

Genetic metrics, discussed further in the Genetic Metric Calculations section, are calculated at two levels for the user defined number of neutral variants per sample at the two ways genomes are tracked in the model by ancestry (roots) and genotype (alleles). The levels are 1) “Effective” modeled truth measurement which identifies unique strains by both ancestry and genotype, e.g. removes exact clones that would not be distinguishable in any capacity and 2) “Genotype” measurement which identifies unique strains by genotype alone, e.g. removes duplicate clones by genotype that may have different ancestries but would not be distinguishable by sequencing. Unless otherwise specified, population-level diversity metrics are reported using the genotype representation, since this most closely matches the information available from empirical sequencing data. Simulated IBD-based relatedness metrics are calculated using the modeled effective representation, because these metrics require ancestry information from allele roots. Detailed chromosome coordinates, allele-root inheritance rules, and recombination implementation are provided in S1 Appendix A–B.

### Intra-host infection model

FPG uses EMOD’s existing mechanistic within-host malaria model to simulate parasite dynamics, gametocyte production, clinical symptoms, and acquired immunity. Infections progress from liver-stage parasites to asexual blood-stage replication and sexual differentiation into mature gametocytes. Host responses include density-dependent innate inflammatory responses and adaptive antibody responses to parasite antigens, including MSP and PfEMP1 variants. These within-host dynamics determine parasite densities, infectiousness to mosquitoes, and whether infections are classified as symptomatic in the observational sampling model. Full details of the EMOD within-host model are described in [23].

FPG extends this within-host model by tracking each genetically distinct parasite strain separately within an infected individual. Each strain carries its own genome and contributes to the host’s circulating parasite and gametocyte populations. As a result, within-host genetic diversity can arise through cotransmission, when multiple genomes are transmitted in a single mosquito bite, or through superinfection, when additional genetically distinct infections are acquired over time.

The relationship between parasite genome and PfEMP1 repertoire is configurable. In the present study, each strain is assigned a random PfEMP1 repertoire, rather than inheriting antigenic repertoires through recombination or imposing age-structured antigenic associations. Thus, the simulations retain EMOD’s immune-mediated infection dynamics and clinical outcome model, but do not assume that genetically related parasites necessarily express more similar PfEMP1 repertoires, or that infections in children and adults are systematically enriched for different PfEMP1 repertoires beyond what emerges from host immune history. This simplifying assumption avoids imposing poorly characterized rules about antigenic inheritance, but may affect simulated within-host competition, clinical presentation, and the relationship between symptomatic sampling and transmission.

### Mosquito-stage transmission and recombination

FPG replaces EMOD’s aggregate contagion-pool representation with explicit tracking of the parasite genomes involved in each human–vector transmission event. As in base EMOD, mosquitoes select human hosts for blood meals according to individual biting risk, age-dependent body surface area, heterogeneous biting preferences, and intervention effects such as bed net use. If a mosquito successfully feeds on an infectious individual, FPG samples mature male and female gametocytes from that host’s circulating gametocyte pool according to their densities and source genomes.

Successful human-to-vector transmission produces one or more oocysts in the mosquito midgut. For each oocyst, one male and one female gametocyte are selected to pair. If the paired gametocytes have identical genomes, the resulting oocyst produces clonal progeny. If the paired gametocytes are genetically distinct, meiosis is simulated to generate four recombinant offspring genomes. Recombination follows an obligate-chiasma model calibrated to *P. falciparum* laboratory crosses, with one obligate crossover per chromosome and additional secondary crossovers placed according to a gamma-distributed inter-crossover distance.

Oocyst maturation is modeled as a time- and temperature-dependent process, after which sporozoites are released and stored as genotype-specific cohorts in the mosquito. Mosquitoes can therefore carry multiple sporozoite genotypes generated from multiple oocysts or blood meals. During a subsequent infectious bite, transmitted sporozoites are sampled from the mosquito’s sporozoite cohorts in proportion to their abundance. If one or more sporozoites successfully establishes liver-stage infection, the corresponding parasite genomes initiate new human infections.

This structure allows FPG to represent genetically diverse infectious bites, distinguish cotransmission from superinfection, track clonal propagation and recombinant lineages, and reconstruct the parasite genomes involved in each transmission event. Parameterization of oocyst burden, oocyst maturation, sporozoite production, sporozoite mortality, hepatocyte invasion, and the full recombination algorithm are provided in S1 Appendix A–B.

### Model calibration

We calibrated 12 EMOD parameters governing within-host parasite dynamics, transmission, and acquired immunity against age-stratified clinical incidence and parasite prevalence data from three reference sites spanning a wide transmission gradient. Calibration followed a two-stage history-matching approach. Candidate parameter sets were first filtered for plausibility against site-level annual entomological inoculation rate (EIR), then evaluated against age-stratified incidence and prevalence likelihoods.

The calibration was intended to ground the epidemiological behavior of the model before adding parasite genetic analyses. Genetic data were not used as calibration targets. As a result, the simulations presented here should be interpreted as mechanistic explorations of how genetic metrics respond under calibrated epidemiological dynamics, rather than as site-specific fits to observed parasite genetic datasets. Full details of calibrated parameters, prior bounds, reference sites, emulator-based filtering, and likelihood calculations are provided in S1 Appendix C.

### Model scenarios

The simulated transmission conditions reflect a seasonal (Sahelian) setting with a single vector species. Seasonality was imposed using a monthly spline of available vector habitat, with transmission peaking in August and September. Transmission intensity varied across scenarios by scaling the overall amplitude of habitat availability. For all scenarios in this study, parasite genomes carried a 100-locus biallelic neutral SNP barcode, with loci distributed across the 14 *P. falciparum* chromosomes approximately in proportion to chromosome length.

Burn-in simulations were used to establish population immunity and endemic parasite diversity across transmission intensities. Each burn-in simulation began with an immunologically naïve population of 10,000 individuals and was run for 50 years, with vital dynamics configured to maintain an approximately sub-Saharan African age distribution. Case management was introduced during the final decade of burn-in and continued throughout pickup simulations. Treatment was provided to 60% of uncomplicated clinical cases in children under 5 years and 30% of uncomplicated clinical cases in individuals 5 years and older; severe cases were treated in 90% of children under 5 years and 80% of individuals 5 years and older.

Pickup simulations combined two serialized burn-in populations into a two-node source–sink model. Source and sink nodes could have different transmission intensities, and importation into the sink node arose through round-trip human movement between nodes. Each individual traveled from their home node to the other node at a fixed per-capita trip rate, spent an exponentially distributed duration away with mean 30 days, and then returned home. Across simulations, the annual trip rate ranged from 0.01 to 1 trip per person per year. There was no importation from outside the modeled two-node system. At the start of pickup simulations, allele frequencies were reset to defined node-specific values while preserving as much of the burn-in parasite population structure as possible; allele roots were also reset so that ancestry-based metrics reflected transmission during the pickup period. Pickup simulations are run for 6 years.

For the matched low- and high-importation intervention comparison shown in Figure 3B, we selected source–sink simulations with similar pre-intervention sink-node incidence but different importation ratios. A single mass drug administration campaign was applied in the sink node, with 90% demographic coverage. We compared post-intervention incidence trajectories to assess how continued importation altered the apparent durability of local intervention response.

We used insecticide-treated net distribution as the primary intervention scenario. ITNs were distributed in the intervention arm on June 1 of year 0 at 80% coverage, and simulations were analyzed over the period spanning three years before through three years after distribution. Net killing efficacy began at 60% and decayed exponentially with a 4-year time constant, while blocking efficacy began at 90% and decayed with a 2-year time constant. Net use varied seasonally with the transmission season and by age, with individuals aged 5-20 years less likely to use a net if they had one. Nets were discarded over time with a median retention time of approximately 5 years, and newborns received nets throughout the simulation period. Intervention simulations were compared with matched no-intervention counterfactual simulations.

For analyses comparing importation regimes, we summarized importation pressure in the sink node as the proportion of new sink-node infections attributable to parasites originating from the source node. Low- and high-importation scenarios were then compared to assess how continued parasite movement altered the relationship between local incidence and parasite genetic metrics.

### Observational sampling model

To mimic field parasite genetic data collection, genetic metrics were calculated from samples drawn from the total simulated infected population. Infected individuals were eligible for sampling at most once per month, so persistent infections could not contribute multiple samples within the same monthly sampling period. Monthly genetic metrics were calculated from eligible infections in each month.

For yearly analyses, samples were drawn from the monthly eligible infection pool under alternative sampling schemes. We varied three features of sampling design: annual sample size, sampling window, and clinical (symptomatic) composition of sampled infections. For sample-size analyses, yearly samples of *N*_infections_were drawn at specified sample sizes. If fewer eligible infections were available than the requested sample size for a given stratum, all available eligible infections were included.

Temporal sampling windows were defined relative to the seasonal incidence peak. windows are defined as: “Peak transmission season” (3-month window centered on the month of highest clinical incidence, October through December), “Full transmission season” (5-month window spanning two months before and after peak incidence, August through January of the following year), and “Full year” (12-month window). For each simulation and sampling scheme, three independent sampling replicates were drawn to capture within-simulation sampling variability.

For clinical-composition analyses, eligible infections were stratified by symptomatic status and sampled as symptomatic infections, asymptomatic infections, or all infections. Additional optional sampling schemes and subpopulation comparisons, including age stratification, monogenomic sampling bias, equal monthly sampling, and source-sink sampling proportions, are described in S1 Appendix D.

### Genetic metric calculations

Genetic metrics were calculated from the sampled parasite genomes at individual and population levels (Table 1, Figure 1E). Metrics based on within-host diversity, including complexity of infection (COI), polygenomic proportion, within-host relatedness, and within-host fixation index (F_WS_), were summarized across sampled individuals. Population-level metrics, including allele frequency, expected heterozygosity, unique genome proportion, and population relatedness, were calculated across genomes in the sampled population.

**Table 1:** EMOD FPG genetic metric calculation descriptions.

| Genetic metric | Description | Calculation | Scale<sup>1</sup> |
| --- | --- | --- | --- |
| Allele frequency per position | The proportion of alternative alleles in all genomes of sampled infections | $\frac{\text{Number of alternative alleles}}{\text{Total alleles}}$ | Population |
| Complexity of infection (COI)<sup>2</sup> | Modeled effective COI: The number of unique genomes after comparing the ancestry of genomes in a sampled individual. | $\sum \text{Unique}(N_{\text{EMOD genome ID}})$ | Individual, summarized by population |
| | Genotype effective COI: The number of unique genomes from a genotype representation of $N_{\text{variants}}$ | $\frac{\sum (N_{\text{infections}}, \text{COI} > 1)}{N_{\text{infections}}, \text{COI} > 1}$ | Individual, summarized by population |
| Cotransmission proportion | The proportion of polygenomic $N_{\text{infections}}$ that contain a bite originating from two different mosquito vectors | $\frac{\sum (N_{\text{infections}}, \text{COI} > 1, \text{Unique}(\text{Bite IDs} > 1))}{N_{\text{infections}}, \text{COI} > 1}$ | Individual, summarized by population |
| Simulated identity by descent (simIBD)<sup>3</sup> | Compare pairwise similarity between two genomes by Hamming distance (H). Uses the initialized root genotype representation, where each ancestor in the simulation is represented at all $N_{\text{variant}}$ positions by an integer. | $\text{IBD}(i, j) = \frac{1 - H(i, j)}{N_{\text{variants}}}$<br>IBD is summarized for all unique pairs of i, j in an individual or population | Within polygenomic individuals and across all genomes in a sampled population group |
| Genotype identity by state (IBS) | Same as IBD, but uses the genotype representation for $N_{\text{variants}}$ | $\text{IBS}(i, j) = \frac{1 - H(i, j)}{N_{\text{variants}}}$ | Individual and population |
| Expected heterozygosity per position ($H_e$) | The probability that two randomly selected genomes carry distinct alleles at a position | $H_e = 1 - (p^2 + q^2)$<br>where P is the reference allele and q is the alternative allele (1-P) | Within polygenomic individuals and across all genomes in a sampled population group |
| Within host fixation index ($F_{ws}$)<sup>4</sup> | Measures the genetic diversity of parasite genomes within one person ($H_w$) compared to the whole population ($H_s$) | $F_{ws} = 1 - \beta$<br>where $\beta$ is the slope of regressing $H_w$ against mean $H_s$ across minor allele frequency bins | Individual, summarized by population |
| Polygenomic proportion<sup>5</sup> | The proportion of $N_{\text{infections}}$ in a sampled population that have a COI > 1 | $\frac{\sum (N_{\text{infections}}, \text{COI} > 1)}{N_{\text{infections}}}$<br>Monogenomic proportion is calculated for both modeled and observed COI | Population |
| Intrahost relatedness ($R_H$)<sup>6</sup> | Co-transmission or super infection inference method comparing within polygenomic sample barcode heterozygosity to expected population monogenomic pair heterozygosity. | $R_H = \frac{H_{\text{mono}} - H_{\text{poly}}}{H_{\text{mono}}}$<br>where $H_{\text{mono}}$ is a random draw from the distribution of monogenomic infection IBS and $H_{\text{poly}}$ is the barcode heterozygosity (number of mixed genotype calls). $R_H$ calculations are repeated for 200 draws per infection. | Individual, summarized by population |
| Unique genomes<sup>5</sup> | All genomes | $\frac{\text{Number of unique genomes, all infections}}{\text{Total genomes in all infections}}$ | Population |
| | Monogenomic infections only | $\frac{\text{Number of unique genotypes in COI = 1 infections}}{\text{Total genomes in all COI = 1 infections}}$ | Population |

<sup>1</sup> Sampled group population level and individual level summary statistics include the number of pairwise comparisons, mean, median, standard deviation, 25<sup>th</sup> and 75<sup>th</sup> quartiles, minimum, and maximum values.
<sup>2</sup> The expected behavior of these metrics is modeled effective COI >= genotype effective COI. The differences between the COI metrics will vary based on epidemiology and the population allele frequency.
<sup>3</sup> Simulated IBD cannot be directly compared to inferred IBD from empirical data. Simulated IBD does not account for any relatedness at the roots; pairwise relatedness will start at zero and increase throughout the simulation. However, the rate and magnitude of pairwise relatedness can be compared to empirical data.
<sup>4</sup> $F_{ws}$ binning concept obtained from moimix R package[https://github.com/bahlolab/moimix]. [24]
<sup>5</sup> Proportion polygenomic and unique genomes are calculated for each level of genome representation as COI - effective modeled truth and by genotype uniqueness within individuals as the strain representatives for population level comparisons.
<sup>6</sup> $R_H$ is included as a calculation but only suggested for 24-position barcodes.

Metrics were calculated using the modeled effective and genotype representations described above. Simulated identity-by-descent metrics were calculated using allele roots from the modeled effective representation, while identity-by-state metrics were calculated from shared allele values.

Monthly metrics were calculated for sampled infections at each monthly time point. Pairwise relatedness metrics, including simulated IBD and IBS, were calculated at the yearly scale for computational tractability. For yearly analyses, metrics were calculated after applying the specified observational sampling scheme, allowing comparisons across sample size, sampling window, and clinical-composition assumptions. Full metric definitions, formulas, and reporting scales are provided in Table 1.

### Simulation outputs and analysis

FPG produces outputs at three levels: individual infection states, transmission events, and observationally sampled infections. Individual-level outputs record host demographics, clinical status, parasite and gametocyte densities, and parasite genomes at user-specified time points. Transmission-event outputs record the source human, biting vector, transmitted parasite genomes, and parental gametocyte genomes, enabling reconstruction of transmission histories and distinction between cotransmission and superinfection. Observational sampling outputs record the subset of infections selected under specified sampling schemes in a format aligned with empirical parasite genetic datasets. These outputs were used to calculate genetic metrics from sampled infections and compare them with simulated clinical incidence and parasite prevalence.

All components of the EMOD with FPG pipeline can be accessed at https://github.com/EMOD-Hub/.

### Generative AI statement

OpenAI ChatGPT (GPT-5.4 and GPT-5.5) and Anthropic Claude Opus (versions 4.5-4.7) were used to assist with code development and to proofread the manuscript for clarity, grammar, and consistency. These tools were used to improve efficiency and readability, not to replace authorial, scientific, or analytical judgment. All AI-generated suggestions, text edits, and code were critically reviewed, revised where necessary, and verified by the authors, who take full responsibility for the accuracy, integrity, and final content of the manuscript.

## Results

### Genetic metrics detect ITN-driven shifts in transmission

We first asked whether parasite genetic metrics could detect an intervention-driven perturbation in transmission under conditions where the epidemiological impact of the intervention was known. To do this, we simulated high-coverage insecticide-treated net (ITN) distribution in a strongly seasonal Sahelian setting and compared genetic and epidemiological trajectories against a no-intervention counterfactual.

In the low-transmission stratum, corresponding to approximately 100–250 clinical cases per 1000 individuals per year, ITN distribution produced clear reductions in both clinical incidence and parasite prevalence immediately after deployment, followed by a gradual return toward baseline transmission over approximately three years (Figure 2A). Clinical incidence and prevalence were correlated across the year, but the shape of this relationship varied across the transmission season (Figure 2B). In particular, the same incidence value could correspond to different prevalence values depending on whether transmission was rising or falling within the seasonal cycle. This seasonal dependence was also observed across transmission strata, with greater seasonal variation at higher transmission intensities (Supplemental Figure 2.1A,B).

**Figure 2.**
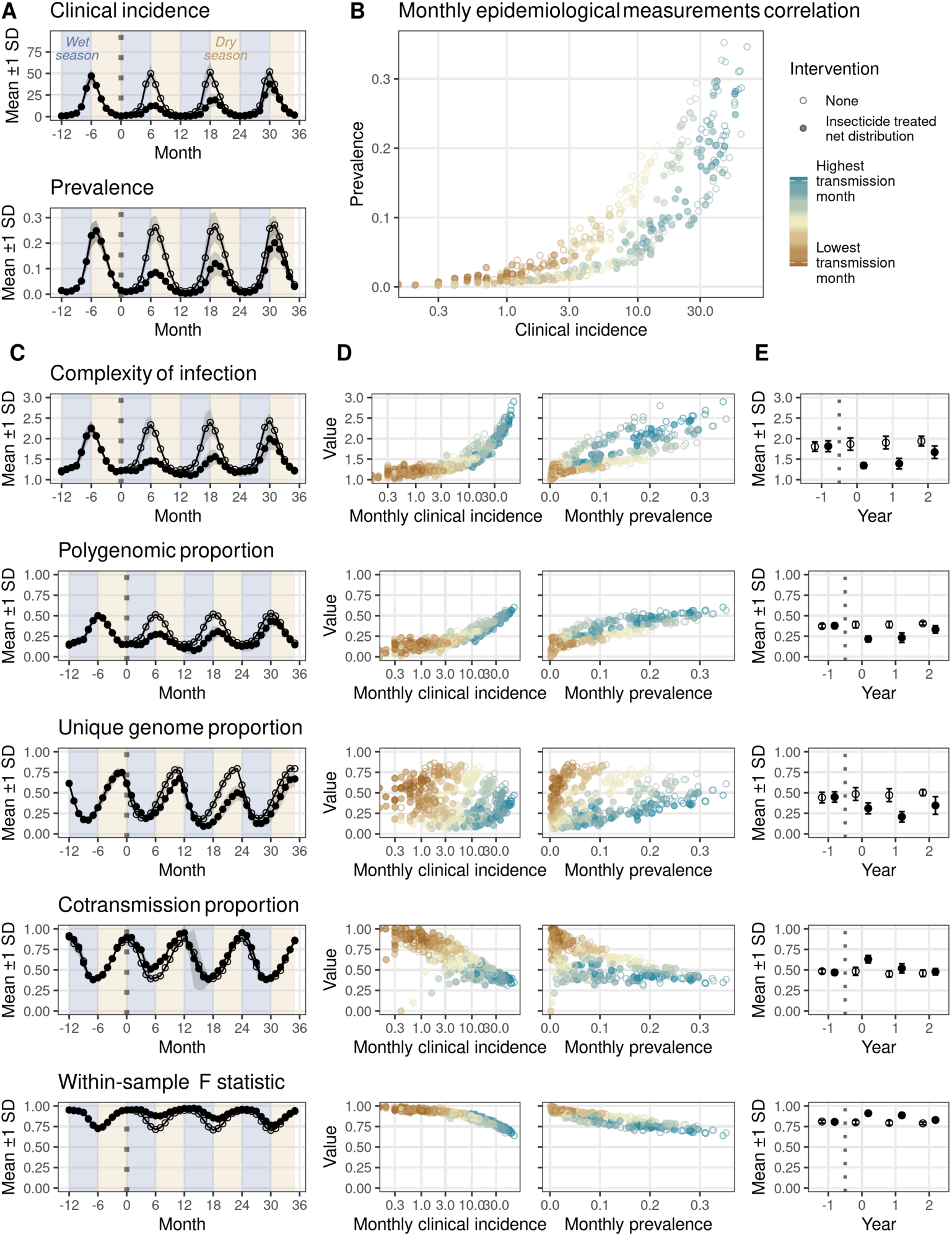
ITN distribution produces shift in genetic metrics across monthly and yearly scales in a low transmission Sahelian seasonality scenario (100-250 cases per 1000 individuals per year). (A) Monthly time series of clinical incidence and prevalence with and without ITN distribution. ITNs are distributed at month 0 (vertical dotted line). (B) The monthly relationship between clinical incidence and prevalence varies across the transmission season. (C) Monthly time series of genetic metrics, corresponding to the same scenarios as shown in Panel (A) and (D) the relationship with each genetic metric per month to clinical incidence and prevalence. (E) Yearly genetic metric trends by ITN distribution calculated from 1000 randomly sampled infections per year. The yearly genetic metrics are sensitive to the changes in transmission caused by the ITN distribution.

Genetic metrics exhibited cyclical dynamics that tracked the wet and dry seasons, with a roughly three-month lag relative to peak transmission (Figure 2C). The ITN distribution causes a sustained change across genetic metrics compared to the no-intervention control scenario, with the divergence becoming apparent within the first seasonal cycle and persisting until the transmission level returns to its pre-intervention baseline after about three years (Figure 2C). The association of monthly genetic metrics and epidemiological measurements varied by metric (Figure 2D). Metrics such as complexity of infection showed stronger monthly correlation with prevalence than with incidence, consistent with their sensitivity to cumulative infection rather than acute transmission events. In contrast, the proportion of unique genomes had weak correlations with clinical incidence but showed clustering by month. Seasonal trends and variance were observed across transmission strata, with smaller seasonal changes for the very low transmission stratum (Supplemental Figure 2.1 C, D).

At the yearly scale, point estimates with uncertainty derived from 1000 randomly sampled infections per year confirmed that intervention-driven reductions in genetic metrics are detectable (Figure 2E), suggesting that population-level genetic signals can reflect intervention impact without requiring exhaustive sampling. The yearly changes in genetic metrics are observed across transmission strata, with a stronger relationship between genetic and epidemiological metrics using yearly measurements rather than monthly (Supplemental Figure 2.2).

### Importation rate can decouple parasite genetic metrics from local transmission intensity

Parasite genomic surveillance for non-resistance applications often assumes that genetic metrics have a stable relationship with local transmission intensity. However, field studies have shown that this relationship can break down in low-transmission settings, where infections may be disproportionately introduced from elsewhere rather than generated by local onward transmission [7,9]. We therefore used the two-node source-sink model to test how varying importation pressure changes the association between parasite genetic metrics and local clinical incidence.

When importation rates were similar across simulations, yearly genetic metrics showed strong relationships with clinical incidence across the broader transmission gradient. However, allowing importation rates to vary introduced substantial noise into these relationships, especially at low incidence (Figure 3A; Supplemental Figure 3.1). The effect of importation differed by metric. Metrics that summarize within-host or population-level parasite diversity, including COI, polygenomic proportion, and unique genome proportion, showed the strongest evidence of an inverse relationship at very low transmission: below approximately 10 clinical cases per 1000 individuals per year, these metrics could be elevated relative to settings with higher incidence. This pattern was most apparent under higher importation pressure, where low local incidence did not necessarily imply low parasite diversity. Thus, low-incidence settings with substantial importation could resemble higher-transmission settings genetically despite limited local transmission.

**Figure 3:**
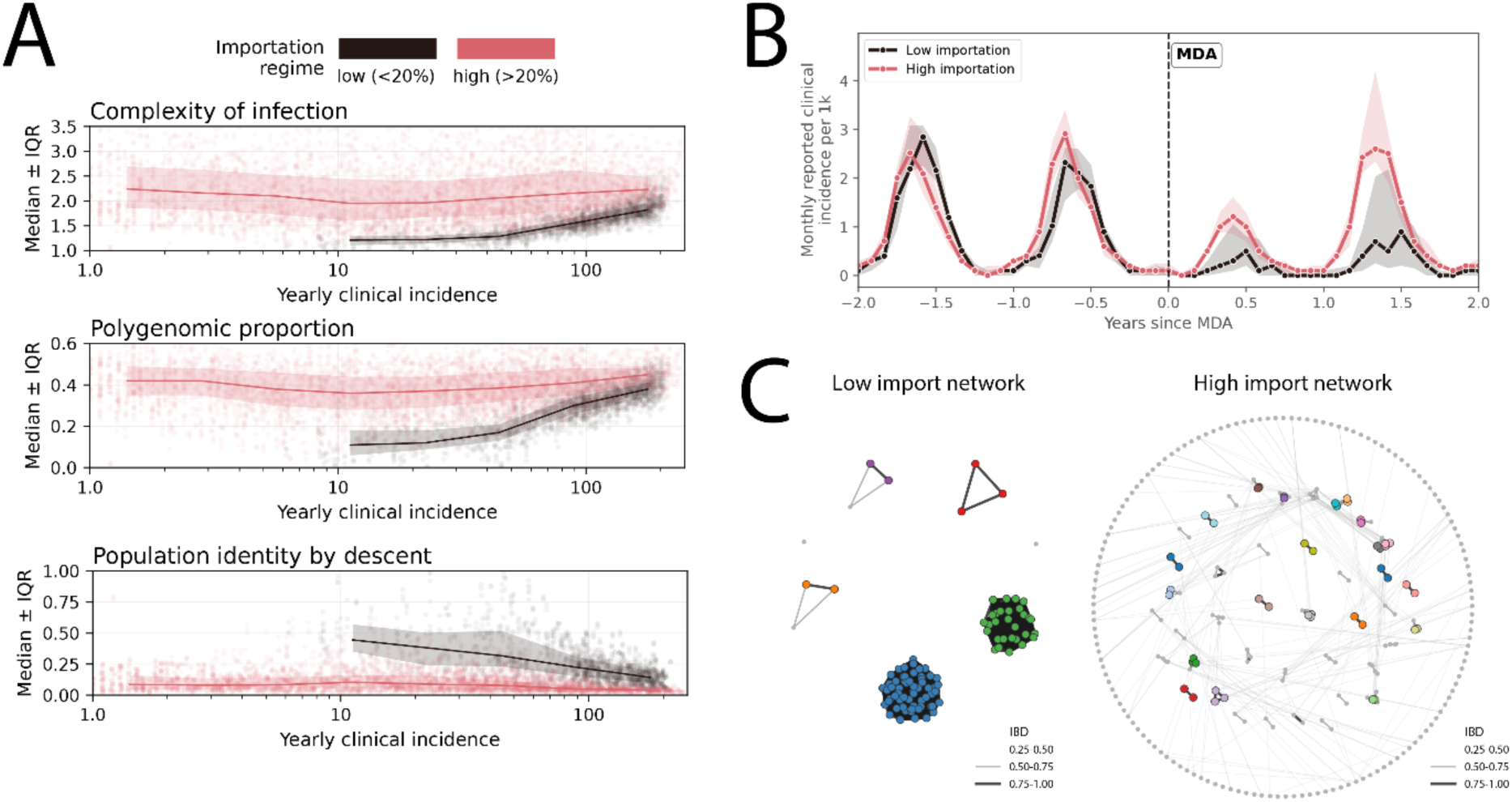
High importation can decouple genetic metrics from local transmission intensity (A) Yearly genetic metrics in the sink population across simulated transmission and importation scenarios, stratified by importation regime. Low importation is defined as <20% of new sink-node infections originating from the source population, and high importation as >20%. Points show simulation-year estimates; lines summarize trends by importation regime. High-importation settings show elevated or less incidence-dependent genetic diversity at low local incidence, weakening the expected relationship between local transmission intensity and parasite genetic metrics. (B) Monthly clinical incidence trajectories for matched low- and high-importation scenarios with similar pre-intervention incidence. A mass drug administration (MDA) campaign is applied at the indicated time point. Despite comparable baseline incidence, high-importation settings show a weaker and less durable response, consistent with continued reintroduction of infections from the source population. (C) Simulated ancestry-IBD connectivity plots for one matched low- and high-importation pair from panel B. One hundred infections were sampled in a single year, and each point represents an individual parasite genome. Colors indicate unique clone groups, defined by exact root identity across the 100 tracked barcode positions. The low-importation network is dominated by a small number of clonal lineages, whereas the high-importation network contains more unique clones and a less locally clustered genetic structure.

In contrast, relatedness-based metrics showed a different pattern. Population identity by descent (IBD) and identity by state (IBS) were less tightly coupled to clinical incidence across the low-transmission range when importation varied. In low-importation settings, low incidence was more often associated with locally clustered or clonal parasite populations consistent with limited onward transmission from a small number of lineages. High-importation settings introduced additional parasite lineages into the sink population, reducing the apparent signal of local clonal expansion and dampening the relationship between relatedness metrics and incidence (Figure 3A; Supplemental Figure 3.1). Within-sample relatedness, IBS, IBD and F_WS_, were comparatively more invariant across the lowest transmission strata, suggesting that some relatedness-based summaries may be less sensitive to local incidence alone when importation pressure is heterogeneous.

This decoupling was also evident in matched low- and high-importation scenarios with similar baseline incidence. After the simulated mass drug administration intervention shown in Figure 3B, high-importation settings showed a weaker and less durable reduction in incidence than matched low-importation settings, consistent with continued reintroduction of infections from the source population. The corresponding ancestry-IBD networks illustrate the same mechanism at the parasite level. The low-importation network was dominated by a small number of clonal lineages, whereas the high-importation network contained more distinct clone groups and less local clustering (Figure 3C).

The effect of importation also depended on the epidemiological comparator. The inverse relationship observed for some diversity metrics at very low clinical incidence was not observed when yearly prevalence was used as the comparator (Supplemental Figure 3.1). Together, these results suggest that importation can drive metric-specific deviations from expected genetic metric–transmission relationships. In operational settings, unexpectedly high COI, polygenomic proportion, or unique genome proportion in a low-incidence setting may indicate parasite mobility or importation, while relatedness metrics may help distinguish locally clustered transmission from repeated introductions. However, the ability to detect importation from genetic and epidemiological metrics alone depends on the comparison populations, the metric selected, and the transmission measure used. As a result, mismatches between local incidence and parasite genetic diversity should be interpreted as potential importation signals that require supporting information on human mobility or parasite connectivity.

### Sampling design affects parasite genetic metric estimation

We next examined how common features of field sampling influence parasite genetic metric estimates. Specifically, we evaluated the effects of sample size, collection timing, and the clinical composition of the sampled population across simulated transmission intensities (Figure 4).

**Figure 4:**
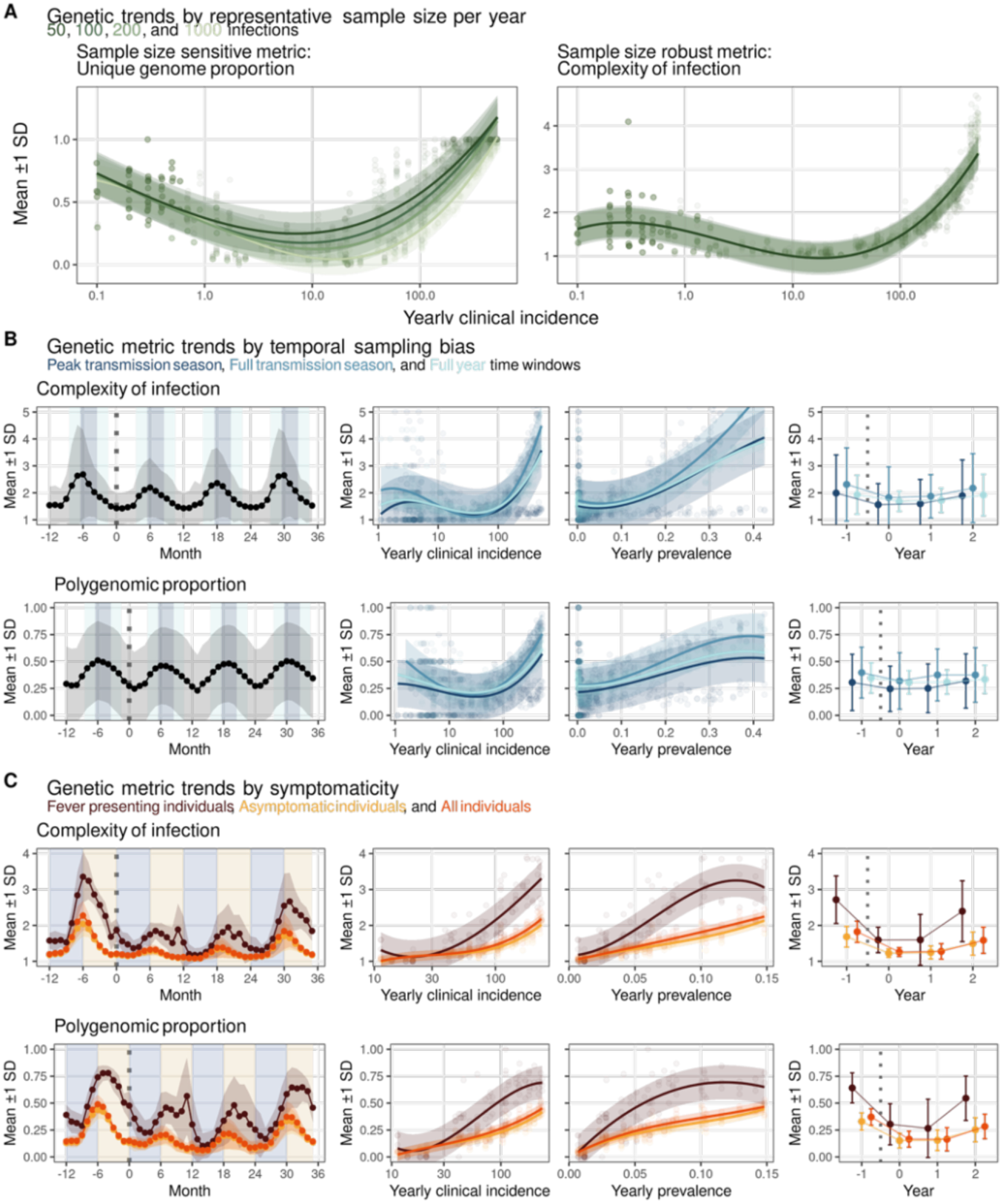
Sampling design affects parasite genetic metric estimates and their relationship to epidemiological measurements. Genetic metrics were recalculated under alternative sampling assumptions to evaluate the effects of sample size, collection timing, and clinical composition of sampled infections. (A) Effect of sample size on genetic metric estimation. Two illustrative metrics are shown: unique genome proportion, which is sensitive to sample size, and complexity of infection, which is comparatively robust. Each line represents a yearly sample size stratum of 50, 100, 200, or 1000 infections. Relationships with yearly clinical incidence are shown across transmission intensities. Additional metrics are shown in Supplemental Figure 4.1. (B) Effect of sampling window on yearly genetic metric estimates and trends in a low transmission stratum. For each genetic metric, three panels show: seasonal trends across years, the cross-sectional relationship between yearly genetic metrics and clinical incidence and prevalence and longitudinal year-to-year changes in genetic metrics. Monthly estimates are calculated from all infections, while yearly estimates are derived from 100 randomly sampled infections per year. Sampling windows are defined as: "Peak transmission season" (3-month window centered on the month of highest clinical incidence), "Full transmission season" (5-month window spanning two months before and after peak incidence), and "Full year" (all samples within a 12-month window). Additional metrics are shown in Supplement Figure 4.2. (C) Effect of individual symptomaticity on genetic metric estimates and trends in a low transmission stratum. Fever-presenting individuals represent ∼11% of the simulated infected population and show elevated genetic metric values and temporally offset seasonal peaks relative to asymptomatic individuals. For each genetic metric, three panels show: seasonal trends across years by symptomaticity group, cross-sectional relationships with yearly clinical incidence and prevalence, and longitudinal year-to-year changes by symptomaticity group. Monthly estimates are calculated from all infections, while yearly estimates are derived from 100 randomly sampled infections per year stratified by symptomaticity. Additional metrics are shown in Supplement Figure 4.3.

Sample size affected some genetic metrics more than others. Unique genome proportion was strongly sample-size dependent when calculated from the 100-position genotype representation (Figure 4A). At a fixed transmission level, unique genome proportion decreased as the number of sampled infections increased, because larger samples created more opportunities to observe repeated clonal genomes in the population. This sample-size dependence was evident across transmission strata and indicates that unique genome proportion cannot be compared directly across datasets with different sample sizes without adjustment. By contrast, complexity of infection and several other relatedness metrics were comparatively robust to sample size, with estimates stabilizing even at sample sizes of approximately 50 infections in many scenarios (Figure 4A; Supplemental Figure 4.1).

Sampling window, or timing, also affected genetic metric estimates, although the effect varied by metric and outcome comparison. Restricting samples to peak transmission season, the broader transmission season, or the full year produced similar cross-sectional relationships between yearly genetic metrics and clinical incidence or prevalence for many metrics (Figure 4B; Supplemental Figure 4.2). However, the sampling window influenced the absolute values of some genetic metrics and the slope of longitudinal year-to-year changes. Peak-season sampling produced higher variance than broader seasonal or year-round sampling windows, reflecting the stronger influence of short-term seasonal dynamics when samples were collected over a narrower interval.

The clinical composition of the sampled population produced the largest systematic differences in genetic metric estimates. Symptomatic individuals had consistently higher genetic metric values than asymptomatic individuals or the full infected population, and their seasonal peaks were temporally offset (Figure 4C; Supplemental Figure 4.3). Fever-presenting individuals represented only approximately 11% of infected individuals in these simulations, so sampling only symptomatic cases selected a small and genetically distinct subset of the infected population. These differences were visible both in monthly seasonal trajectories and in yearly summaries stratified by symptomaticity.

Together, these results show that sampling design can alter the estimated magnitude and variability of parasite genetic metrics. Sample size was especially important for metrics based on the proportion of unique genomes, sampling window affected seasonal and longitudinal estimates, and symptomaticity introduced systematic differences between passively sampled clinical infections and the broader infected population.

## Discussion

Malaria molecular surveillance is increasingly being considered for programmatic use cases beyond monitoring drug resistance and diagnostic marker deletions, including assessing transmission changes, evaluating intervention impact, and identifying importation. However, interpreting parasite genetic metrics for these broader use cases requires understanding when they reflect local transmission and when they are shaped by external processes such as importation or sampling design. Here, we presented EMOD with Full Parasite Genetics (FPG), a mechanistic simulation framework that links malaria transmission, within-host infection dynamics, mosquito-stage recombination, parasite genetic diversity, and observational sampling. Using seasonal simulations across transmission intensities, we evaluated how common parasite genetic metrics respond to intervention, importation, and sampling processes relevant to malaria molecular surveillance.

Our first major finding was that parasite genetic metrics can detect intervention-driven changes in transmission under controlled sampling conditions. In a seasonal Sahelian setting, ITN distribution produced reductions in clinical incidence and prevalence that were accompanied by detectable shifts in multiple genetic metrics at both monthly and yearly timescales. These changes emerged within the first post-intervention transmission season and persisted while epidemiological impact remained detectable. However, the relationship between genetic and epidemiological measurements was strongly metric-specific. COI and polygenomic proportion were more closely aligned with prevalence than with clinical incidence. Other metrics, such as unique genome proportion, were more sensitive to seasonal timing and population bottlenecks. These results suggest that parasite genetic metrics can reflect intervention impact, but that metric choice and sampling timing strongly influence signal strength.

Our second major finding was that importation can decouple parasite genetic metrics from local transmission intensity, especially at very low incidence. In low-importation settings, declining incidence was often associated with locally clustered or clonal parasite populations. In contrast, high-importation settings could maintain elevated COI, polygenomic proportion, or unique genome proportion despite low local incidence, because imported infections introduced additional parasite lineages into the sink population. This pattern provides a mechanistic explanation for empirical observations in low-transmission settings such as Senegal, Sao Tome and Príncipe, Zambia, and parts of Uganda where genetic diversity does not decline monotonically with incidence [7,9,25,26]. Operationally, mismatches between low clinical incidence and unexpectedly high parasite genetic diversity may therefore be useful as a flag for parasite mobility or importation. However, these mismatches should not be interpreted as definitive evidence of importation without supporting mobility, connectivity, or epidemiological data.

Our third major finding was that sampling design can substantially alter genetic metric estimates. Sample size had especially strong effects on unique genome proportion, because larger samples increased the probability of observing repeated clonal genomes. Sampling window influenced absolute metric values, temporal trends, and variance, particularly when samples were restricted to peak transmission season. The clinical composition of sampled infections produced the largest systematic differences: symptomatic infections had higher genetic metric values and temporally shifted seasonal peaks relative to asymptomatic infections and the full infected population. These results indicate that passive case-based sampling and active population-based sampling may not produce directly comparable genetic summaries, even within the same simulated transmission setting.

Taken together, these results show that parasite genetic metrics are sensitive to intervention- and importation-driven changes in transmission, but that this sensitivity is conditioned by how samples are collected and how metrics are selected and interpreted. The three-month seasonal lag and the differential responsiveness of metrics to incidence or prevalence mean that even well-designed surveillance programs need to be deliberate about timing and metric choice. At the same time, the sample size sensitivity of certain metrics, the effects of restricted collection windows, and the systematic overrepresentation of symptomatic individuals in passive surveillance each introduce biases capable of masking or mimicking real epidemiological change.

The epidemiological indicator used as the benchmark for genetic metrics also shapes their interpretation. In this study, we focused on clinical incidence and RDT prevalence because these are among the transmission measures most commonly available to NMCPs across diverse settings. However, a broader set of transmission indicators should be explored. Adjusted incidence estimates, which account for variation in care-seeking behavior, access to care, and reporting completeness, may improve the interpretability and generalizability of comparisons with genetic metrics. Serological indicators may provide complementary information by reflecting recent or cumulative exposure, depending on the marker and assay used. EIR remains a direct measure of transmission intensity but is rarely available at the spatial and temporal resolution needed for routine comparison with genetic surveillance data. Exploring how genetic diversity metrics correspond to a broader set of transmission measures and across diverse settings could strengthen the evidence base for MMS as a routine surveillance tool and help identify which genetic metrics are best suited to specific epidemiological contexts.

Modeling can help identify which genetic metrics best balance high information value with strong operational feasibility. Our modeling scenarios suggest that most metrics are sensitive to interventions and importation when large numbers of samples can be collected. However, the robustness of metrics can vary under more modest sampling levels. For example, polygenomic proportion and within-host fixation index (F_WS_) showed strong monthly and yearly correlations to epidemiological measurements, while others such as unique genome proportion showed poor monthly correlation to epidemiological measurements but was sufficient to track intervention changes and cross-sectional stratum differences at the yearly scale (Figure 2, Figure 4A). This heterogeneity in relationship strength and timing suggests that metric selection should be guided by the specific epidemiological use cases.

These results also have implications for the design and interpretation of malaria molecular surveillance systems. Programs that can influence sample collection should prioritize consistent sampling windows, document the clinical composition of sampled infections, and select metrics that are robust to the expected sample size. When surveillance depends on convenience samples, such as passively detected clinical cases, genetic estimates should be interpreted considering the sampling process that generated them. Cross-site comparisons are especially vulnerable to bias when sites differ in sample size, timing, care-seeking behavior, or the fraction of symptomatic infections represented in the dataset. Standardized reporting of sampling metadata would improve the interpretability and comparability of molecular surveillance outputs.

Several limitations should be noted. First, the simulations presented here focus on a highly seasonal Sahelian transmission setting. The qualitative effects of intervention, importation, and sampling are likely relevant more broadly, but the specific quantitative thresholds, lags, and sample-size requirements should not be treated as general rules. Perennial transmission settings, different vector ecologies, different intervention packages, and different importation structures may produce different genetic trajectories. Second, although the model was calibrated to age-stratified incidence and prevalence data, calibration was performed for a limited set of parameters and did not include genetic calibration targets. As a result, uncertainty in the current analysis reflects stochastic simulation and sampling variation more than full parameter uncertainty. Future work should calibrate jointly to longitudinal epidemiological and genetic time series as such datasets become available.

The genetic model also includes simplifying assumptions. The analyses presented here focus on neutral biallelic markers and do not explicitly evaluate selection, strain-specific fitness differences, drug resistance dynamics, or immune-mediated competition between genetically related parasites. The model currently assumes that all parasite strains present within an infection are detectable, whereas real sequencing data are affected by parasite density, within-host strain frequency, assay sensitivity, and bioinformatic thresholds for calling mixed infections. Incorporating technology-specific observation models would improve comparisons between simulated and empirical genetic metrics. In addition, the present simulations assume random PfEMP1 repertoires for each strain, including sibling parasites generated through recombination. This avoids making strong assumptions about antigenic inheritance, but it may limit the model’s ability to capture relationships between genetic relatedness, antigenic similarity, and within-host competition.

Despite these limitations, FPG provides a flexible framework for evaluating when parasite genetic metrics are likely to be informative for malaria surveillance. By explicitly representing transmission, recombination, within-host diversity, importation, and sampling, the model can be used to test surveillance strategies before applying them to field data. The central conclusion of this study is that though parasite genetics are broadly informative about transmission, the utility of each specific metric depends on the epidemiological question, the transmission context, and the sampling design. Defining those conditions is essential if malaria molecular surveillance is to support programmatic decisions that rely on estimating intervention impact, importation, and transmission changes.

## Data Availability

All components of the EMOD with FPG pipeline are available at https://github.com/EMOD-Hub/. Base EMOD FPG can be found in the emodpy-malaria repository, and the observational layer to match sampling schemes can be found in the FPGObservationalModel repository.

## Acknowledgements

We would like to thank collaborators who helped shape and improve the simulation framework and archetypes, including Wesley Wong, Arnau Pujol, Svetlana Titova, Ye Chen, Sharon Chen, and Christopher Lorton. We would also like to thank Caroline Buckee for providing feedback on the manuscript and Mandy Izzo for improvements to the model schematic diagrams.

## Supplement Figures and Tables

**Supplement Table 1:**
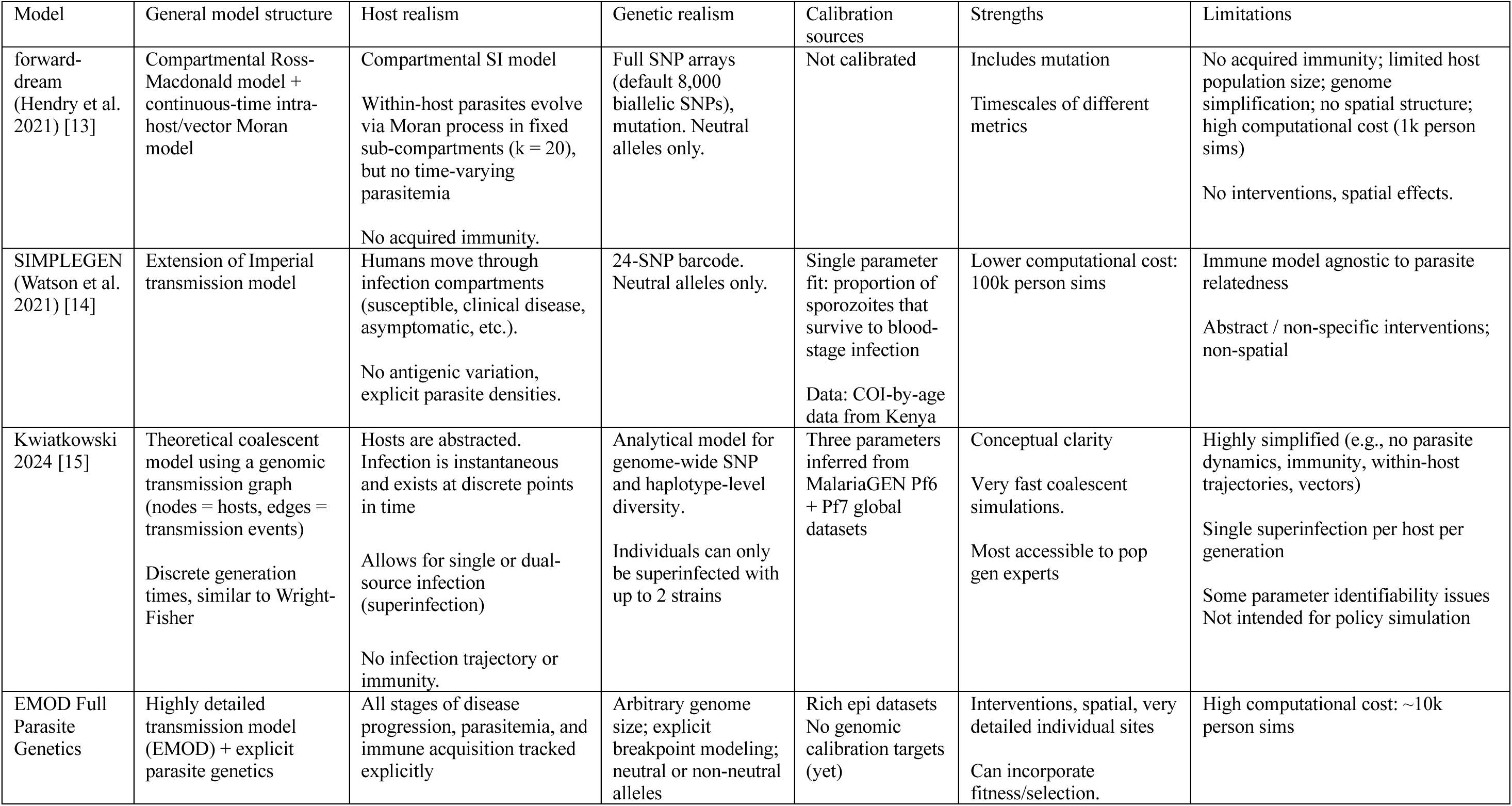
Comparison of available mathematical malaria genetic epidemiology models (neutral alleles only).

**Supplement Figure 1.1:**
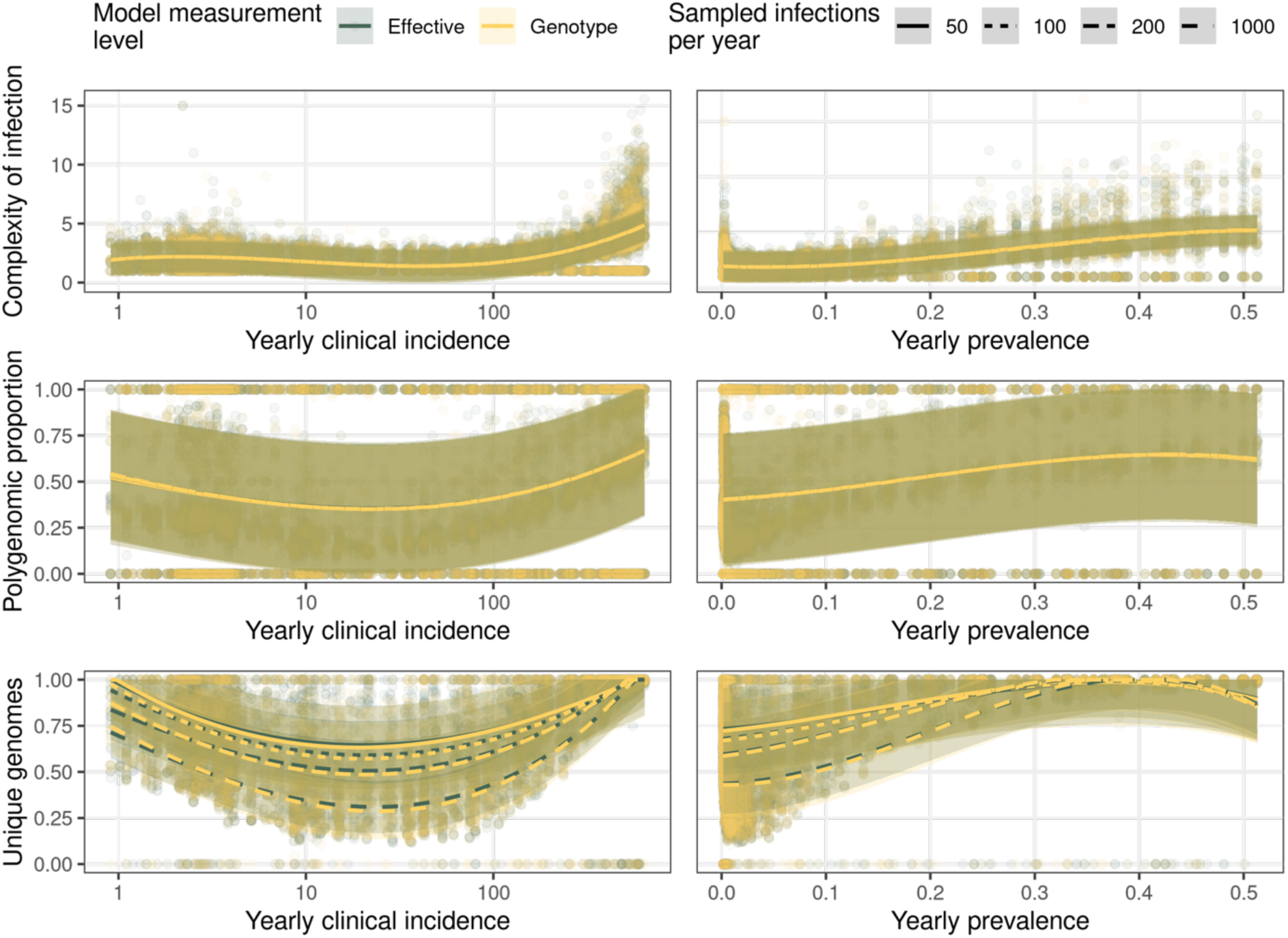
Comparison of modeled effective and genotype representations for selected genetic metrics. Genetic metric relationships with yearly clinical incidence and yearly parasite prevalence are shown using the modeled effective representation and the genotype representation. Metrics are calculated from annual samples of 50, 100, 200, or 1000 infections. In these simulations, the 100-locus barcode was initialized with allele frequencies near 0.5, resulting in similar trends between modeled effective and genotype representations for the metrics shown. This supports the use of the genotype representation for sequencing-like diversity metrics in the main analyses.

**Supplement Figure 1.2:**
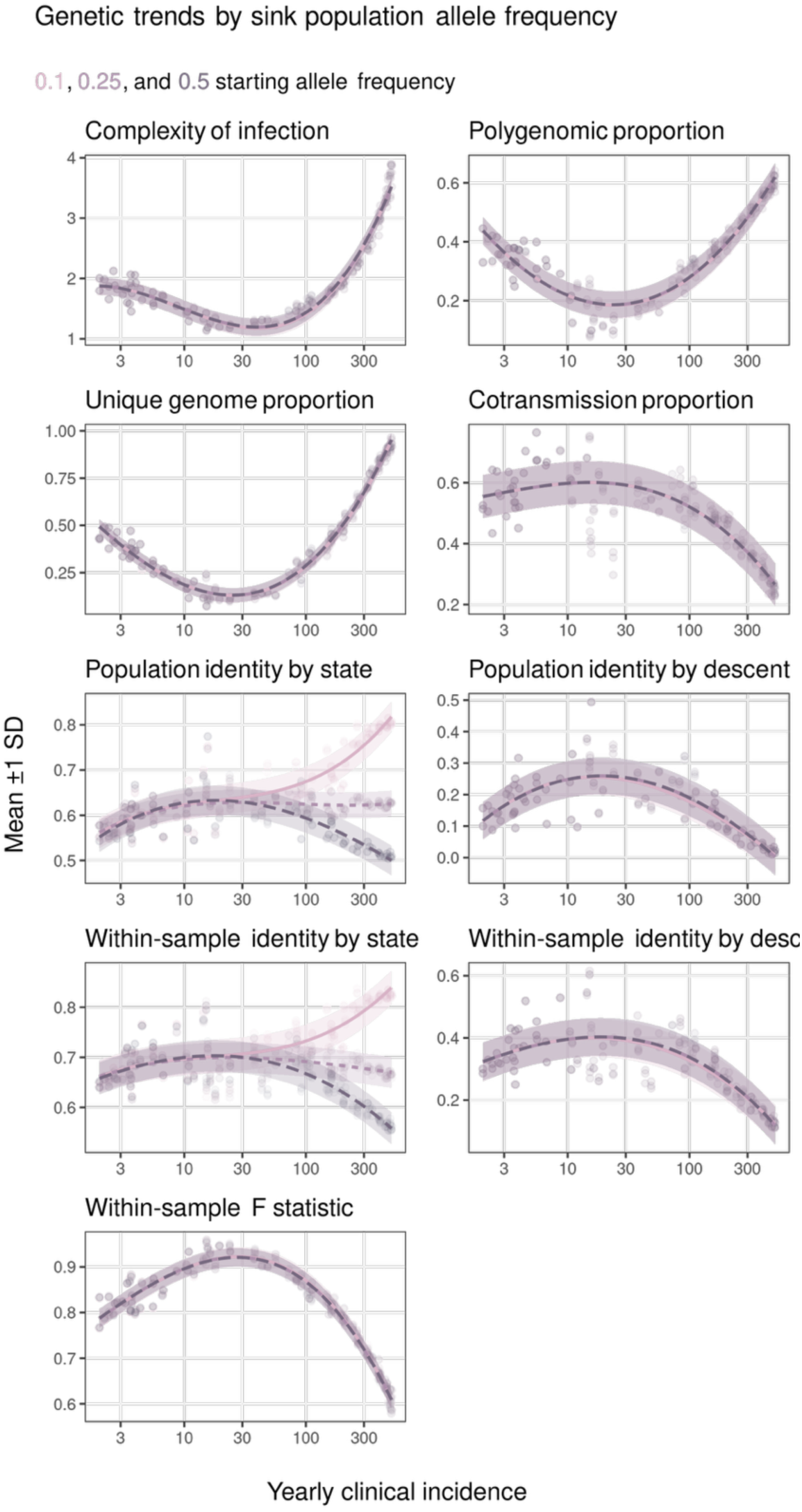
Comparison of model levels on the relationship between genetic metrics and epidemiological measurements for metrics by local (“sink”) population starting allele frequency. Starting population allele frequency has negligible effects on all metrics at very low transmission, with deviations in the slope occurring around the beginning of the low transmission strata (> 100 cases per 1000) for identity by state.

**Supplement Figure 2.1:**
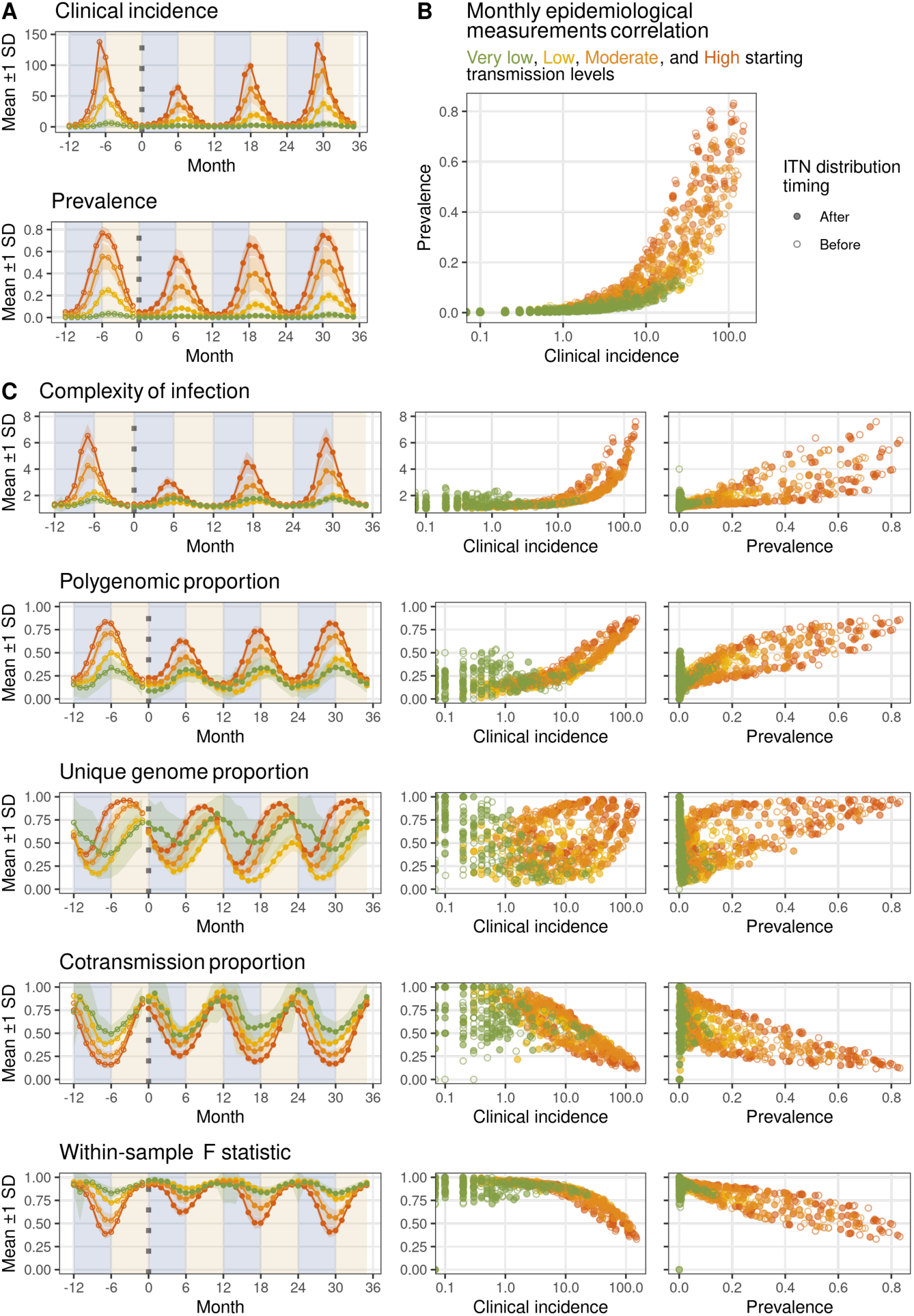
Assessing insecticide treated net (ITN) distribution impact on trends in monthly genetic metrics for WHO transmission categories in a Sahelian seasonality scenario (Very low < 100 (green), low < 250 (yellow), moderate < 450 (orange), high > 450 (red) cases per 1k individuals). (A) Monthly time series in reported incidence and prevalence by WHO transmission categories with an ITN distribution intervention at month 0. (B) Monthly correlation of epidemiological measurements by month colored by seasonality. (C) Monthly trends of genetic metrics and the relationship of metrics to reported incidence and prevalence.

**Supplement Figure 2.2:**
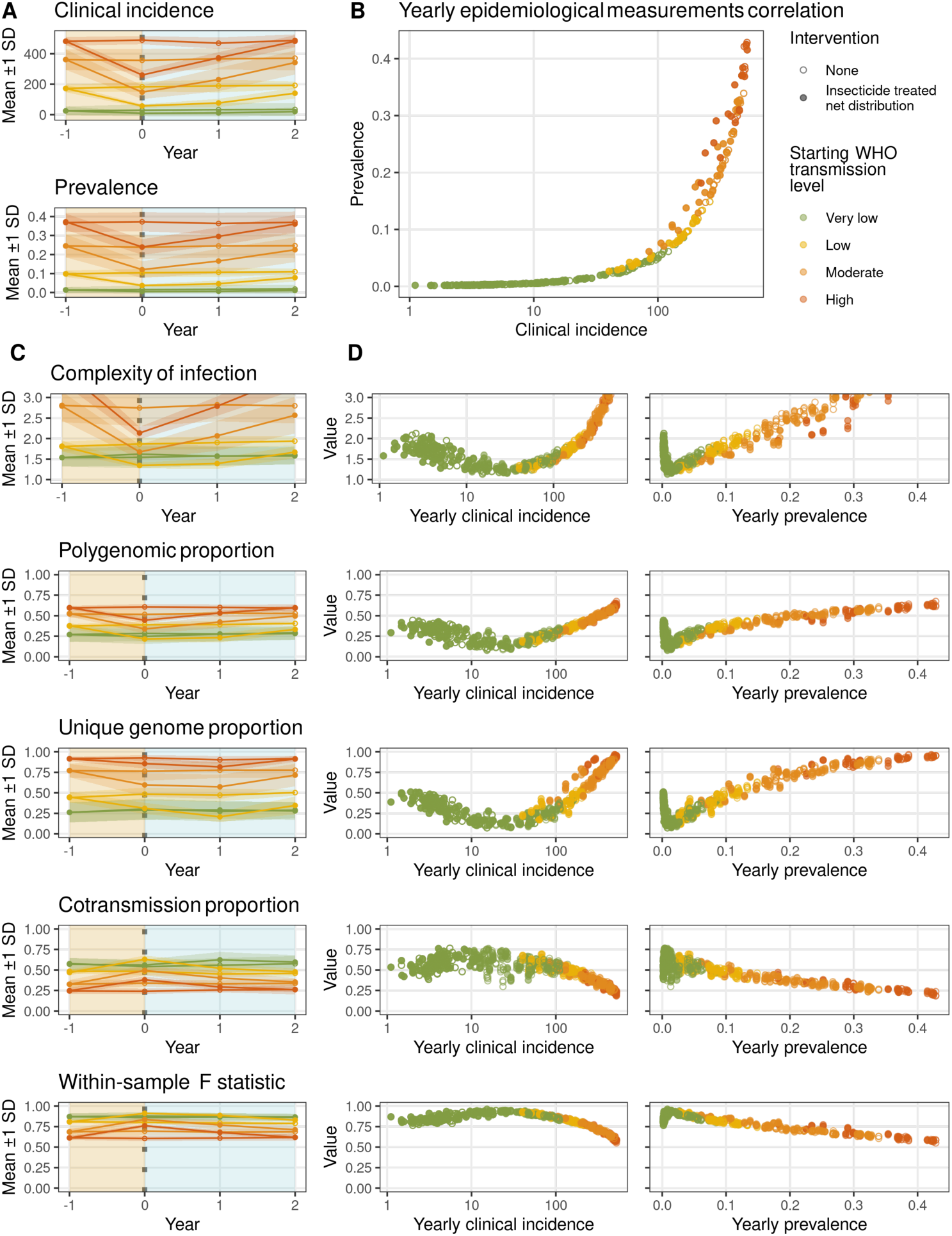
ITN distribution impacts on yearly genetic metric trends across transmission intensity categories in a Sahelian seasonality setting. All yearly metrics are calculated from 1,000 randomly sampled infections per year for three sampling replicates per simulation. (A) Yearly clinical incidence and prevalence time series before and after ITN distribution (month 0) stratified by WHO transmission category: very low (<100 cases per 1000 individuals), low (100–250), moderate (250–450), and high (>450). (B) Cross-sectional relationship between yearly incidence and prevalence, colored by transmission category. (C) Yearly genetic metric trends and their cross-sectional relationships with clinical incidence and prevalence, stratified by transmission category. (D) Yearly genetic metric trends stratified by both transmission category and intervention status (ITN distribution vs. control).

**Supplemental Figure 3.1:**
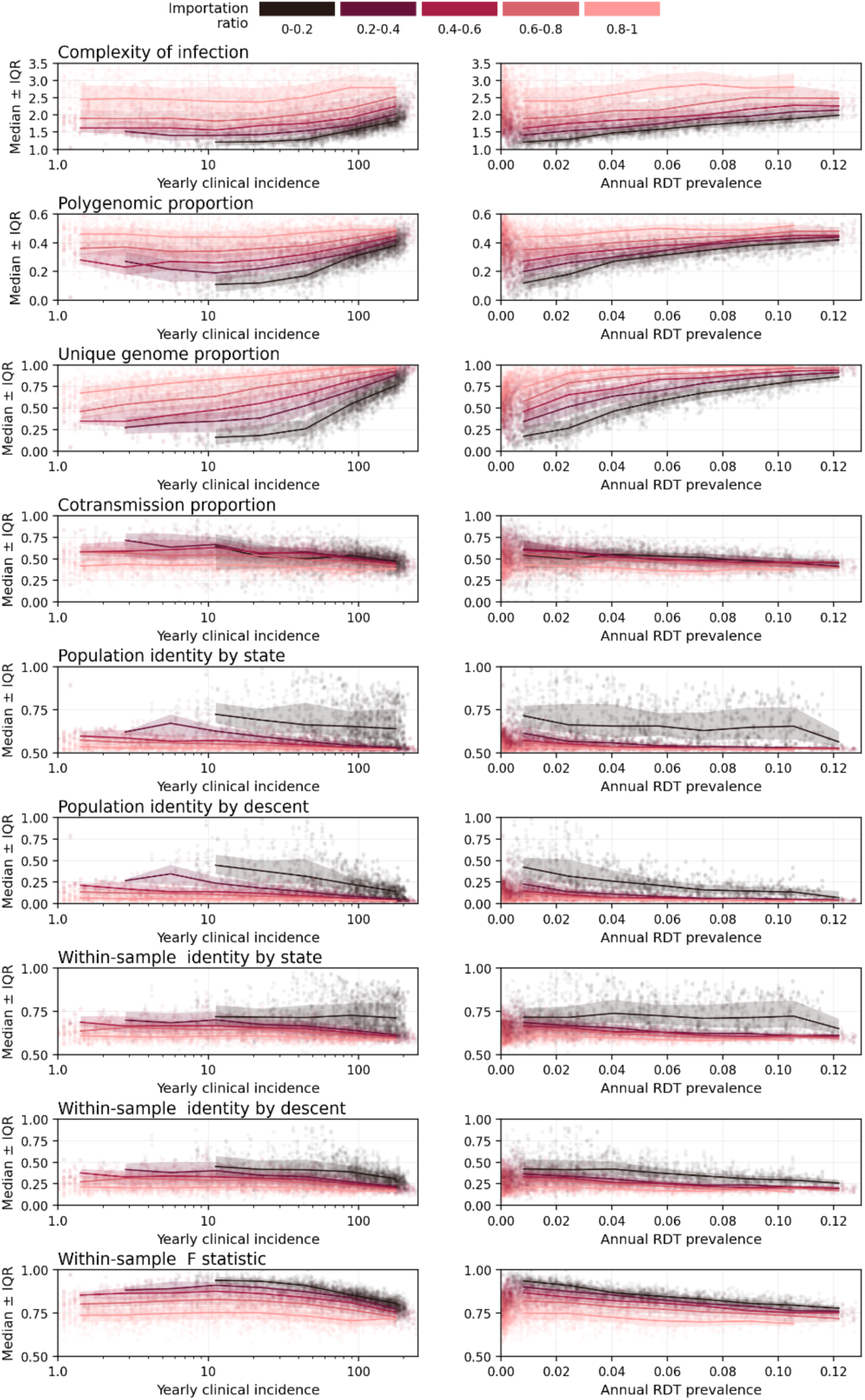
Effects of importation on relationships between parasite genetic metrics and yearly clinical incidence and RDT prevalence. Each point represents three 100-infection random draws per simulation in a single year. Lines and bands show median and interquartile range for given importation level. Importation bins are defined as the fraction of new sink-node infections originating from the source population (darker colors = less importation).

**Supplement Figure 4.1:**
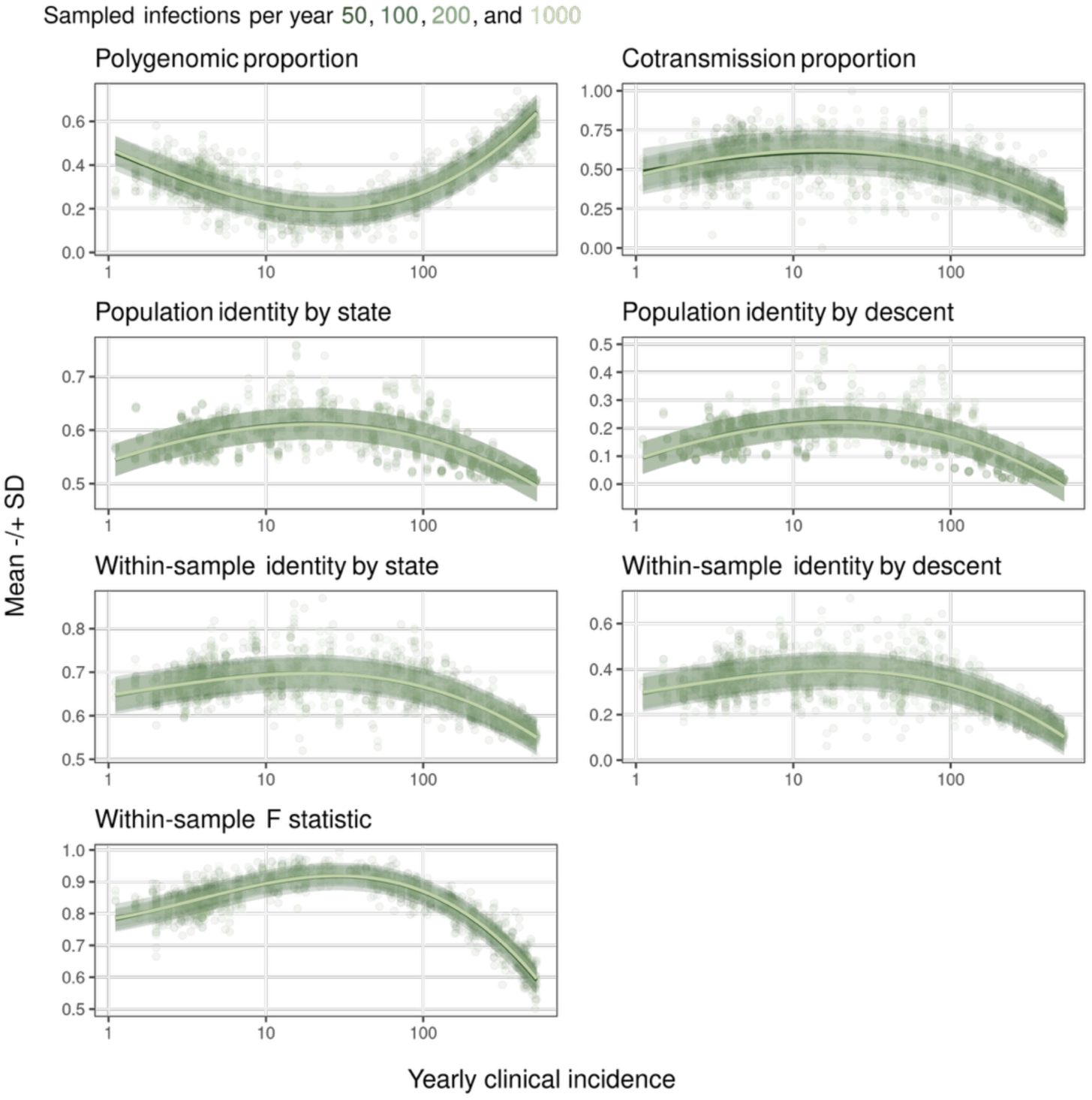
Effect of sample size on genetic metric estimation across transmission strata. Each line represents a distinct sample size stratum (50, 100, 200, or 1000 infections). Relationships with yearly clinical incidence are shown for each stratum.

**Supplement Figure 4.2:**
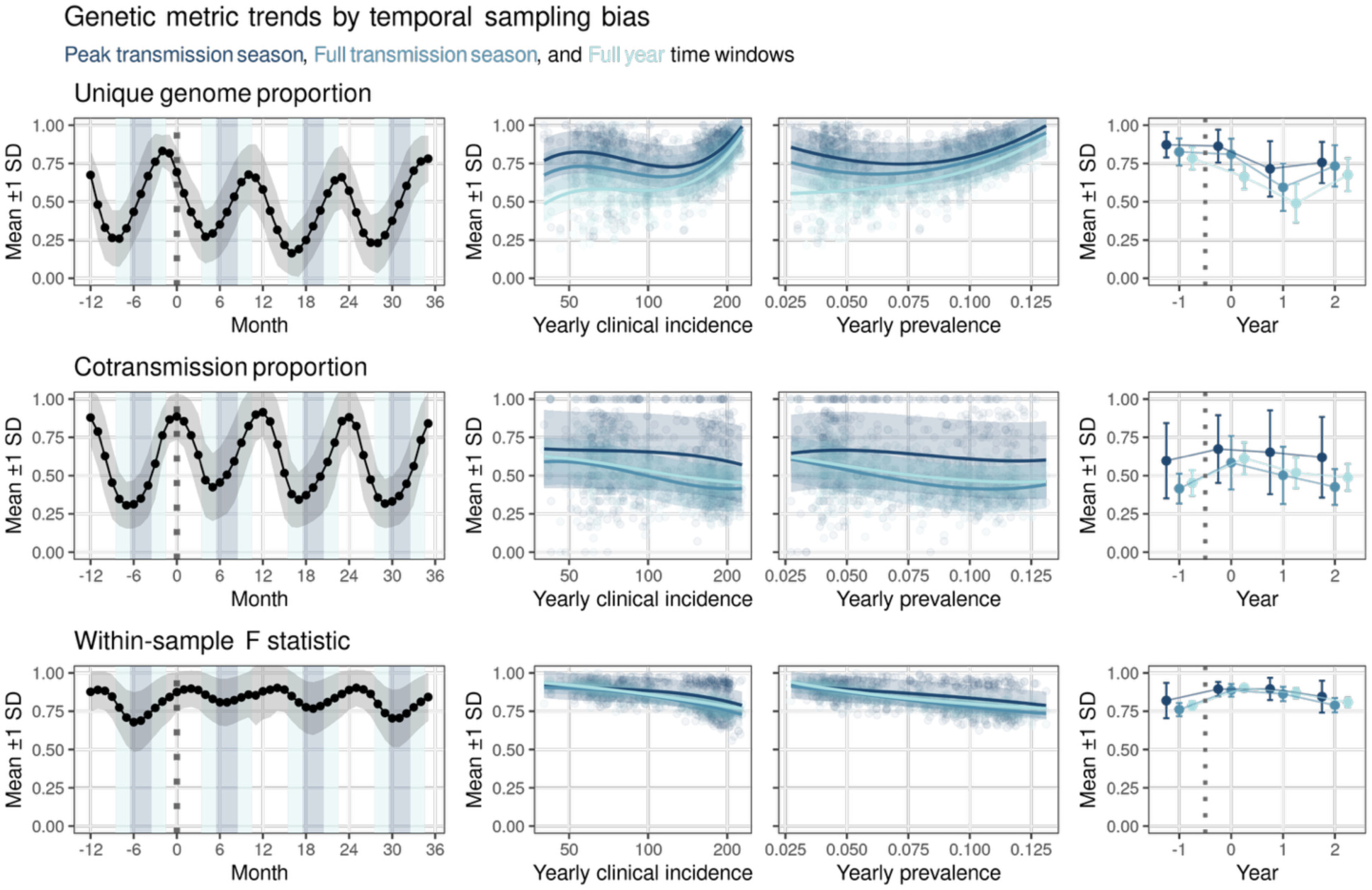
Effect of sampling window on yearly genetic metric estimates and trends in a low transmission stratum. For each genetic metric, three panels show: seasonal trends across years pre and post ITN distribution (month 0, vertical dot line), the cross-sectional relationship between yearly genetic metrics and clinical incidence and prevalence and longitudinal year-to-year changes in genetic metrics. Monthly estimates are calculated from all infections, while yearly estimates are derived from 100 randomly sampled infections per year. Sampling windows are defined as: "Peak transmission season" (3-month window centered on the month of highest clinical incidence), "Full transmission season" (5-month window spanning two months before and after peak incidence), and "Full year" (all samples within a 12-month window).

**Supplement Figure 4.3:**
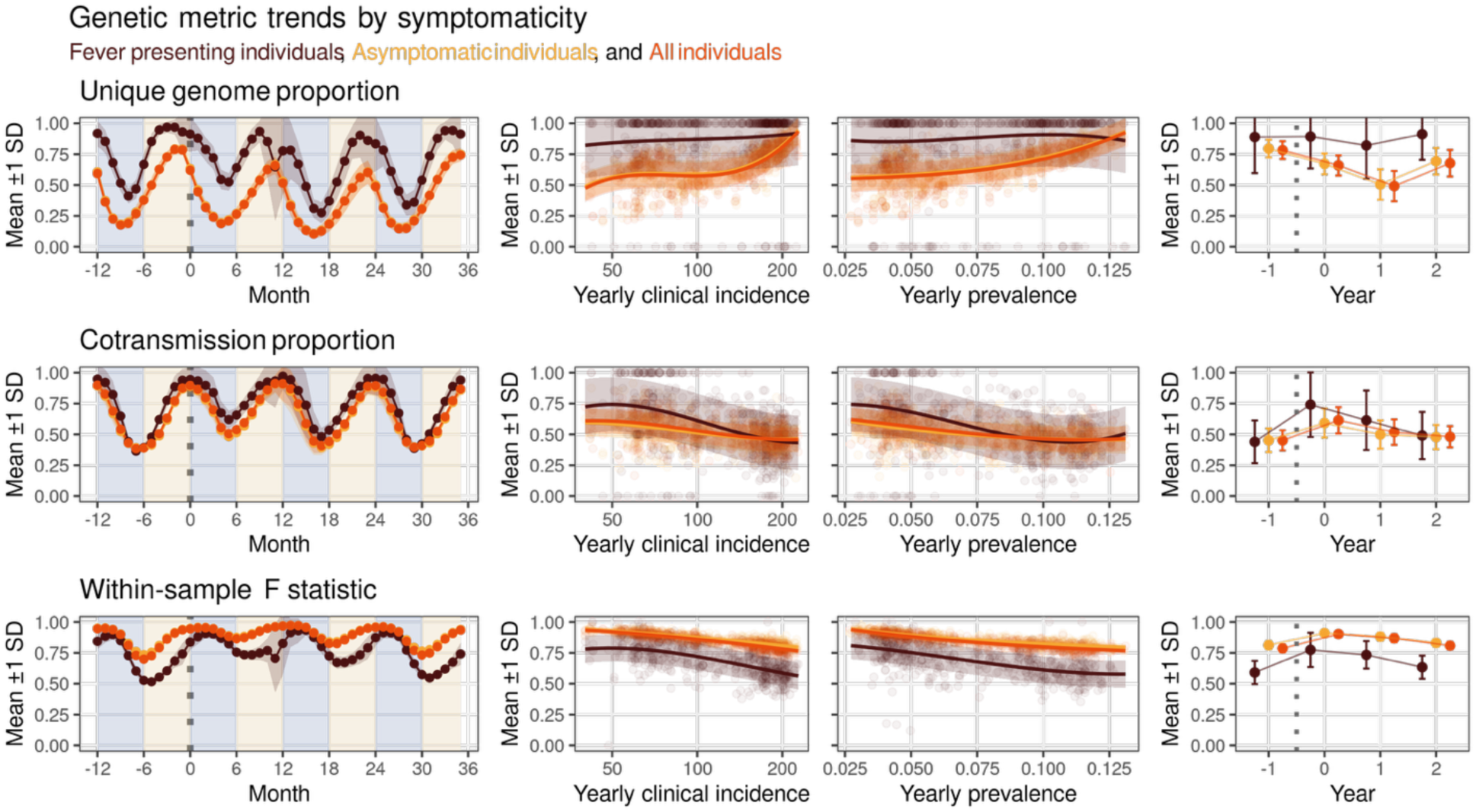
Effect of individual symptomaticity on genetic metric estimates and trends in a low transmission stratum. Fever-presenting show elevated genetic metric values and temporally offset seasonal peaks relative to asymptomatic individuals. For each genetic metric, three panels show: seasonal trends across years pre and post ITN distribution by symptomaticity group, cross-sectional relationships with yearly clinical incidence and prevalence, and longitudinal year-to-year changes by symptomaticity group. Monthly estimates are calculated from all infections, while yearly estimates are derived from 100 randomly sampled infections per year stratified by symptomaticity.

## References

1. Gates Foundation. Malaria Surveillance | Global Grand Challenges. Nov 2020 [cited 22 Apr 2026]. Available: https://gcgh.grandchallenges.org/challenge/new-approaches-integrating-molecular-surveillance-malaria-control-programs-round-26

2. Dada N, Simpson VJ, Amenga-Etego LN, Oriero E, Miotto O, Torok ME, et al. Towards integrated malaria molecular surveillance in Africa. Trends Parasitol. 2024;40: 964–969. doi:10.1016/j.pt.2024.09.005

3. Gatton ML, Dunn J, Chaudhry A, Ciketic S, Cunningham J, Cheng Q. Implications of parasites lacking *Plasmodium falciparum* histidine-rich protein 2 on malaria morbidity and control when rapid diagnostic tests are used for diagnosis. J Infect Dis. 2017;215: 1156–1166. doi:10.1093/INFDIS/JIX094

4. Malaria Policy Advisory Group. Statement by the Malaria Policy Advisory Group on the urgent need to address the high prevalence of pfhrp2/3 gene deletions in the Horn of Africa and beyond. In: Statement by the Malaria Policy Advisory Group [Internet]. 28 May 2021 [cited 6 Apr 2026]. Available: https://www.who.int/news/item/28-05-2021-statement-by-the-malaria-policy-advisory-group-on-the-urgent-need-to-address-the-high-prevalence-of-pfhrp2-3-gene-deletions-in-the-horn-of-africa-and-beyond

5. Moriarty LF, Betoulle MM, Troell P, Cavros I, Griffith K, Plucinski MM. Antimalarial efficacy monitoring after nearly 20 years of artemisinin-based combination therapy in Africa: recalibrating guidance. Trends Parasitol. 2026;0. doi:10.1016/J.PT.2026.02.012

6. WHO Technical Consultation Group. Technical consultation on the role of parasite and anopheline genetics in malaria surveillance. Geneva, Switzerland; 2019 Jul. Available: https://www.who.int/publications/m/item/meeting-report-of-the-technical-consultation-on-the-role-of-parasite-and-anopheline-genetics-in-malaria-surveillance

7. Wong W, Schaffner SF, Thwing J, Seck MC, Gomis J, Diedhiou Y, et al. Evaluating the performance of *Plasmodium falciparum* genetic metrics for inferring National Malaria Control Programme reported incidence in Senegal. Malar J. 2024;23: 68-. doi:10.1186/S12936-024-04897-Z

8. Schaffner SF, Badiane A, Khorgade A, Ndiop M, Gomis J, Wong W, et al. Malaria surveillance reveals parasite relatedness, signatures of selection, and correlates of transmission across Senegal. Nat Commun. 2023;14: 7268-. doi:10.1038/s41467-023-43087-4

9. Kiyaga S, Mbabazi M, Katairo T, Kabbale KD, Asua V, Nsengimaana B, et al. Accuracy of *Plasmodium falciparum* genetic data for estimating parasite prevalence and malaria incidence in Uganda. Malar J. 2026;25: 147-. doi:10.1186/S12936-026-05836-W

10. Verity R, Aydemir O, Brazeau NF, Watson OJ, Hathaway NJ, Mwandagalirwa MK, et al. The impact of antimalarial resistance on the genetic structure of *Plasmodium falciparum* in the DRC. Nat Commun. 2020;11: 2107-. doi:10.1038/s41467-020-15779-8

11. Pujol A, Chidimatembue A, Silva C da, Boene S, Mbeve H, Cisteró P, et al. Estimating probabilities of malaria importation in southern Mozambique through *P. falciparum* genomics and mobility patterns. Elife. 2025;14. doi:10.7554/ELIFE.107136.2

12. Daniels RF, Schaffner SF, Dieye Y, Dieng G, Hainsworth M, Fall FB, et al. Genetic evidence for imported malaria and local transmission in Richard Toll, Senegal. Malar J. 2020;19: 276-. doi:10.1186/S12936-020-03346-X

13. Hendry JA, Kwiatkowski D, McVean G. Elucidating relationships between *P. falciparum* prevalence and measures of genetic diversity with a combined genetic-epidemiological model of malaria. PLoS Comput Biol. 2021;17: e1009287. doi:10.1371/JOURNAL.PCBI.1009287

14. Watson OJ, Okell LC, Hellewell J, Slater HC, Unwin HJT, Omedo I, et al. Evaluating the performance of malaria genetics for inferring changes in transmission intensity using transmission modeling. Mol Biol Evol. 2021;38: 274–289. doi:10.1093/MOLBEV/MSAA225

15. Kwiatkowski D. Modelling transmission dynamics and genomic diversity in a recombining parasite population. Wellcome Open Res. 2024;9. doi:10.12688/WELLCOMEOPENRES.19092.1

16. Eckhoff PA. A malaria transmission-directed model of mosquito life cycle and ecology. Malar J. 2011;10: 303. doi:10.1186/1475-2875-10-303

17. Selvaraj P, Suresh J, Wenger EA, Bever CA, Gerardin J. Reducing malaria burden and accelerating elimination with long-lasting systemic insecticides: a modelling study of three potential use cases. Malar J. 2019;18: 307-. doi:10.1186/S12936-019-2942-4

18. Gerardin J, Bever CA, Bridenbecker D, Hamainza B, Silumbe K, Miller JM, et al. Effectiveness of reactive case detection for malaria elimination in three archetypical transmission settings: A modelling study. Malar J. 2017;16. doi:10.1186/s12936-017-1903-z

19. Gerardin J, Bever CA, Hamainza B, Miller JM, Eckhoff PA, Wenger EA. Optimal population-level infection detection strategies for malaria control and elimination in a spatial model of malaria transmission. PLoS Comput Biol. 2016;12: e1004707. doi:10.1371/journal.pcbi.1004707

20. Nikolov M, Bever CA, Upfill-Brown A, Hamainza B, Miller JM, Eckhoff PA, et al. Malaria elimination campaigns in the Lake Kariba Region of Zambia: A spatial dynamical model. Lloyd-Smith J, editor. PLoS Comput Biol. 2016;12: e1005192. doi:10.1371/journal.pcbi.1005192

21. Wenger EA, Eckhoff PA. A mathematical model of the impact of present and future malaria vaccines. Malar J. 2013;12: 126. Available: http://www.malariajournal.com/content/12/1/126

22. Eckhoff PA. Malaria parasite diversity and transmission intensity affect development of parasitological immunity in a mathematical model. Malar J. 2012;11: 419. doi:10.1186/1475-2875-11-419

23. Eckhoff P. *P. falciparum* infection durations and infectiousness are shaped by antigenic variation and innate and adaptive host immunity in a mathematical model. PLoS One. 2012;7: e44950. doi:10.1371/JOURNAL.PONE.0044950

24. bahlolab. moimix: R package for inferring multiple infections from high-throughput sequencing data GitHub. [cited 23 Apr 2026]. Available: https://github.com/bahlolab/moimix

25. Fola AA, He Q, Xie S, Thimmapuram J, Bhide KP, Dorman J, et al. Genomics reveals heterogeneous *Plasmodium falciparum* transmission and selection signals in Zambia. Communications Medicine. 2024;4: 67-. doi:10.1038/s43856-024-00498-8

26. Chen Y, Ng PY, Garcia-Ruiz D, Elliot A, Palmer B, Assunção Carvalho RMC d’, et al. Genetic surveillance reveals low but sustained malaria transmission with clonal replacement in Sao Tome and Principe. Communications Medicine. 2025;5: 199-. doi:10.1038/s43856-025-00905-8

