## Supplementary material for "EMOD with Full Parasite Genetics: A modeling framework for evaluating parasite genetic metrics for operational malaria molecular surveillance": S1 Appendix

#### Appendix A: Mosquito-stage parameterization and genome tracking

##### A.1 Oocyst development rate

Oocyst maturation in the mosquito midgut is temperature-dependent and follows the Arrhenius relationship used in EMOD's vector model [1]. The daily progress toward sporozoite release is calculated as:

$$\text{progress} = \Delta t \cdot A_1 \cdot \exp\left(-\frac{A_2}{T}\right)$$

where $\Delta t$ is the timestep in days, $T$ is the local air temperature in Kelvin, $A_1 = 1.17 \times 10^{11}$ and $A_2 = 8336$. When cumulative progress reaches 1.0, sporozoites are released.

S1 Appendix Figure 1: Temperature-dependent oocyst development.

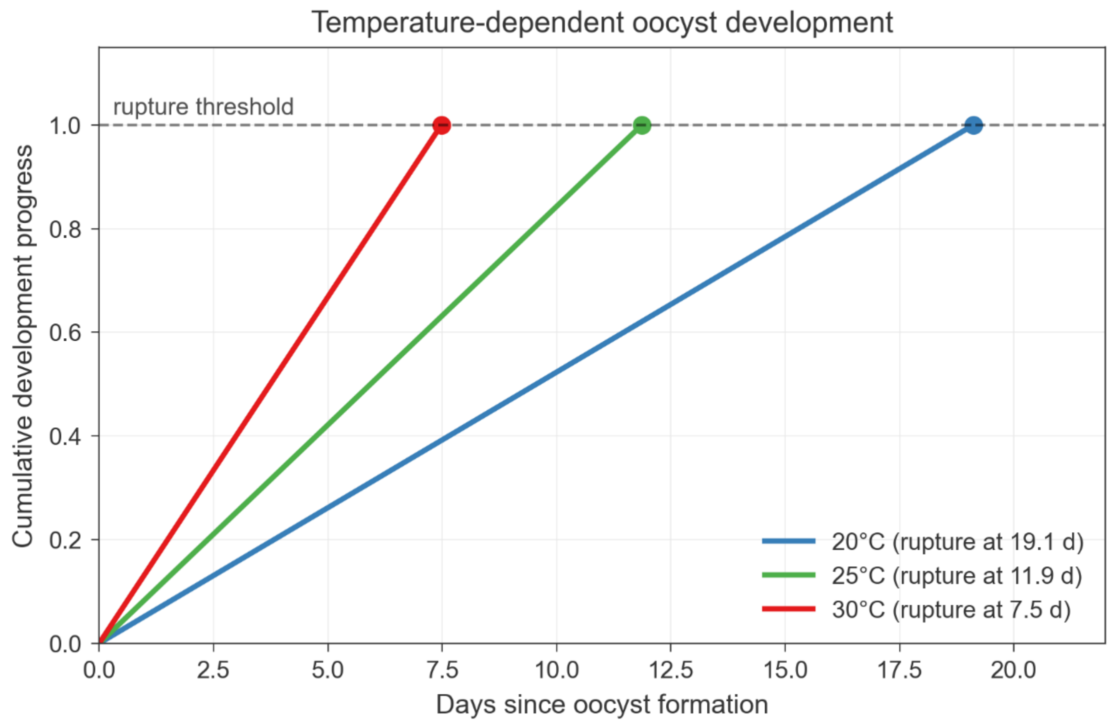


##### A.2 Crossover point distribution

Meiotic recombination follows an obligate chiasma model calibrated to laboratory crosses of *Plasmodium falciparum* [2,3].

##### *Chromosome structure*

The *P. falciparum* genome comprises 14 nuclear chromosomes. The model uses the following chromosome-length values, rounded to the nearest kilobase for internal use and approximating published *P. falciparum* reference lengths [4].

S1 Appendix Table 1: Genomic coordinates of potential crossover locations. Min Location and Max Location define the contiguous genomic coordinate space used for crossover placement.

| **Chromosome** | **Length (bp)** | **Min Location** | **Max Location** |
|---|---|---|---|
| 1 | 643,000 | 1 | 643,000 |
| 2 | 947,000 | 643,001 | 1,590,000 |
| 3 | 1,100,000 | 1,590,001 | 2,690,000 |
| 4 | 1,200,000 | 2,690,001 | 3,890,000 |
| 5 | 1,300,000 | 3,890,001 | 5,190,000 |
| 6 | 1,400,000 | 5,190,001 | 6,590,000 |
| 7 | 1,400,000 | 6,590,001 | 7,990,000 |
| 8 | 1,300,000 | 7,990,001 | 9,290,000 |
| 9 | 1,500,000 | 9,290,001 | 10,790,000 |
| 10 | 1,700,000 | 10,790,001 | 12,490,000 |
| 11 | 2,000,000 | 12,490,001 | 14,490,000 |
| 12 | 2,300,000 | 14,490,001 | 16,790,000 |
| 13 | 2,700,000 | 16,790,001 | 19,490,000 |
| 14 | 3,300,000 | 19,490,001 | 22,790,000 |
| **Total** | 22,790,000 | | |

##### *Obligate chiasma*

Each chromosome receives exactly one obligate crossover at a position drawn uniformly across the chromosome length [2]:

$$x_{\text{obligate}} \sim \text{Uniform}(0, L)$$

where $L$ is the chromosome length in base pairs.

##### *Secondary crossovers*

Additional crossover points are placed bidirectionally from the obligate chiasma. The inter-crossover distance $d$ is drawn from a gamma distribution [2]:

$$d \sim \text{Gamma}(k, \theta)$$

with shape parameter $k = 2$ and scale parameter $\theta = 0.38$ centimorgans (cM). This parameterization yields a mean inter-crossover distance of 0.76 cM, consistent with observed crossover frequencies in *P. falciparum* [2].

##### *Conversion to base pairs*

We follow the parameterization of Wong et al. 2018 [2], in which centimorgan distances drawn from the gamma distribution above are converted to physical distances via:

$$d_{\mathrm{bp}} = d_{\mathrm{cM}} \times 15{,}000 \times 100$$

The 15,000 bp/cM term reflects the empirical recombination density in *P. falciparum* of approximately 15 kilobases per centimorgan [2]. With default parameters, this yields a mean inter-crossover distance of ~1.14 Mb. Secondary crossovers are placed iteratively in both directions from the obligate chiasma until the chromosome boundaries are reached.

#### A.3 Sporozoite and oocyst distributions

##### *Number of oocysts from blood meal*

The number of oocysts that develop in a mosquito following an infectious blood meal follows a zero-truncated negative binomial distribution, calibrated to empirical oocyst burden data [5]. The negative binomial component is parameterized in terms of failures:

$$N_{\mathrm{oocyst}} \sim \mathrm{NegBin}(r, p)$$

where $r$ is the number of failures before stopping and $p$ is the probability of failure per trial. The default parameterization uses $r = 3$ and $p = 0.5$.

S1 Appendix Figure 2: Oocyst distribution per successful blood meal.

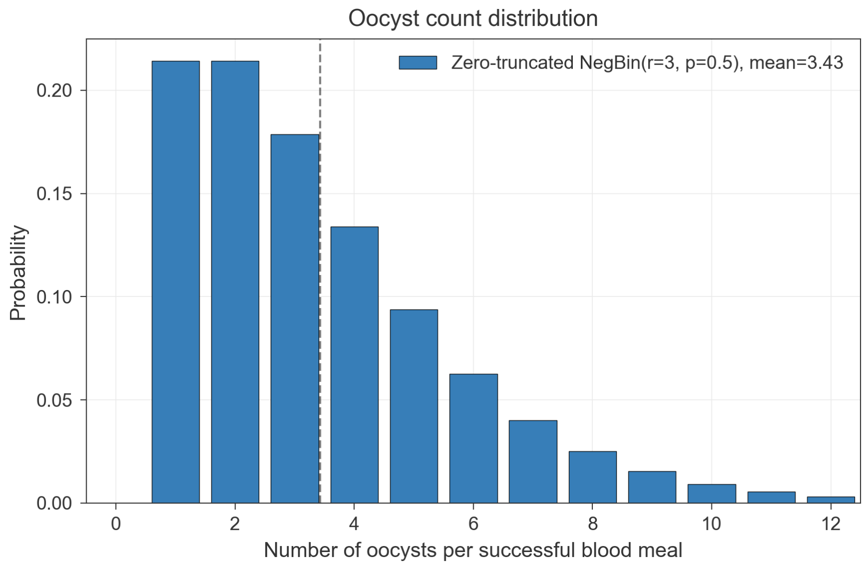


##### *Number of sporozoites per bite*

The number of sporozoites deposited during an infectious bite follows a negative binomial distribution [cite]:

$$N_{\mathrm{spz}} \sim \mathrm{NegBin}(r, p)$$

The default parameterization uses $r = 12$ and $p = 0.5$, yielding an expected sporozoite load consistent with empirical measurements of salivary gland contents [6].

S1 Appendix Figure 3: Per-bite sporozoite distribution.

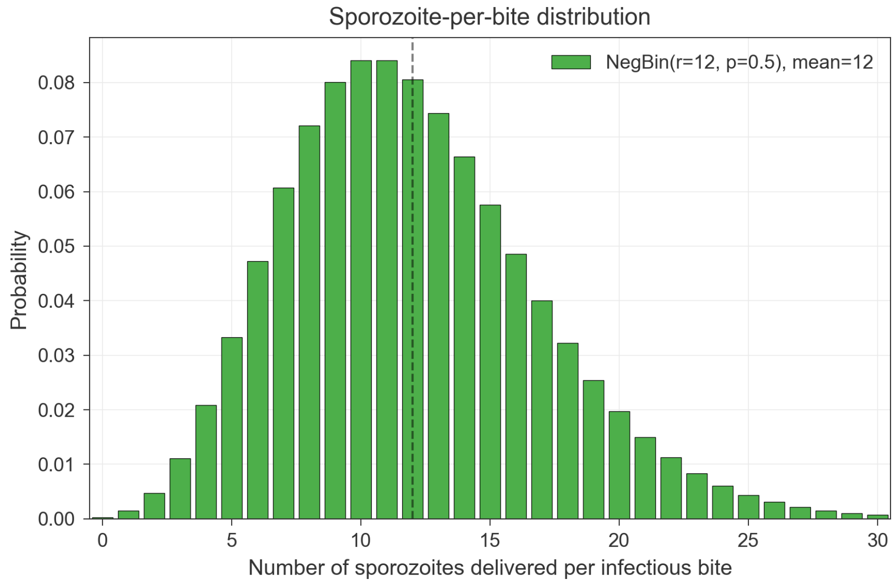

*Number of sporozoites per oocyst*

Upon maturation, each oocyst ruptures to release sporozoites. The number of sporozoites produced per oocyst is drawn from a Gaussian distribution [7]:

$$N_{\mathrm{spz/oocyst}} \sim \mathrm{Normal}(\mu, \sigma)$$

with mean $\mu = 4{,}000$ and standard deviation $\sigma = 1{,}000$.

##### *A.4 Sporozoite mortality*

Sporozoites in mosquito salivary glands can persist after salivary-gland invasion, but empirical studies indicate that their abundance and functional infectivity decline with age. We therefore represent loss of salivary-gland sporozoites using a first-order exponential mortality process [8]. The daily survival probability is:

$$P_{\mathrm{survive}} = \exp\left(-\frac{1}{\tau}\right)$$

where $\tau$ is the sporozoite life expectancy. The default value of $\tau = 10$ days corresponds to a daily mortality rate of approximately 10%. This decay mechanism means that long-lived mosquitoes may become less infectious over time as their sporozoite load diminishes, although new sporozoite cohorts may arise after subsequent infectious blood meals.

#### A.5 Hepatocyte invasion

Not all sporozoites deposited during a bite successfully invade hepatocytes. A baseline survival fraction reflects the inherent inefficiency of sporozoite migration and hepatocyte invasion [cite]:

$$N_{\mathrm{surviving}} = N_{\mathrm{spz}} \times f_{\mathrm{survival}}$$

where we use the default assumption $f_{\mathrm{survival}} = 0.25$.

#### A.6 Identity-by-descent tracking

FPG distinguishes identity-by-state (IBS) from simulated identity-by-descent (IBD) using allele roots. Each parasite genome maintains two parallel arrays: an allele-value array, which stores the genotype at each tracked locus, and an allele-root array, which records the ancestral infection from which each allele descends. When a new infection is introduced exogenously, all allele roots are initialized to the unique identifier of the originating infection, marking that genome as a distinct founding lineage. For infections generated through mosquito transmission, allele roots are inherited from parental gametocyte genomes through the same recombination process as allele values.

During meiotic recombination, allele roots are exchanged together with their corresponding allele values. Thus, if a progeny genome inherits an allele value from a parental chromatid at a given locus, it also inherits that chromatid’s allele root at the same locus. This parallel inheritance allows the model to calculate IBS from shared allele values and simulated IBD from shared allele roots, even after multiple generations of recombination.

#### Appendix B: Meiotic Recombination Algorithm

The algorithm below describes progeny generation for outcrossed oocysts formed from genetically distinct parental gametocytes. When paired male and female gametocytes have identical genomes, recombination is skipped and the oocyst produces clonal progeny.

##### B.1 Overview

When a mosquito ingests gametocytes from an infected blood meal, sexual reproduction occurs in the midgut. The algorithm proceeds in four stages:

1. **Gametocyte pairing**: Male and female gametocytes are paired to form diploid zygotes

2. **Crossover placement**: Recombination breakpoints are determined for each chromosome

3. **Chromatid generation**: Four recombinant chromatids are produced per chromosome

4. **Progeny assembly**: Haploid progeny genomes are assembled through independent assortment

##### B.2 Gametocyte Pairing

The oocyst count is determined before parental genomes are assigned. Specifically, after a successful infectious blood meal, the model draws the target number of oocysts from the zero-truncated negative binomial distribution described in Appendix A.3. Gametocyte pairing is then used to assign one male and one female parental genome to each oocyst. Thus, gametocyte sampling determines the genetic composition of oocysts, and may cap the realized number of oocysts when insufficient male-female gametocyte pairs are available, but does not otherwise generate the initial oocyst count.

```
FUNCTION PairGametocytes(gametocytes_by_genotype_and_sex,
num_oocysts):

    # Count available gametocytes
    male_counts = {genotype: count for each genotype}
    female_counts = {genotype: count for each genotype}

    pairs = []

    FOR i = 1 TO num_oocysts:
        # Sample one male gametocyte (probability proportional to
counts)
```

```
        male_genotype = WeightedSample(male_counts)
        male_counts[male_genotype] -= 1

        # Sample one female gametocyte (probability proportional
to counts)
        female_genotype = WeightedSample(female_counts)
        female_counts[female_genotype] -= 1

        pairs.append((male_genotype, female_genotype))

    RETURN pairs
```

When male and female genotypes differ, the resulting oocyst is outcrossed and will produce recombinant progeny. When they are identical, the oocyst is inbred.

#### B.3 Crossover Placement

For each chromosome, crossover positions are determined using an obligate chiasma model with gamma-distributed secondary crossovers.

```
FUNCTION PlaceCrossovers(chromosome_length, gamma_k,
gamma_theta):

    crossover_positions = []

    # Place obligate chiasma at uniform random position
    obligate_position = Uniform(0, chromosome_length)
    crossover_positions.append(obligate_position)

    # Place secondary crossovers extending left from obligate
    position = obligate_position
    WHILE position > 0:
        distance_cM = Gamma(k=gamma_k, theta=gamma_theta)
        distance_bp = distance_cM * 15000 * 100  # Convert cM to
bp
        position = position - distance_bp
        IF position > 0:
            crossover_positions.append(position)

    # Place secondary crossovers extending right from obligate
    position = obligate_position
```

```
    WHILE position < chromosome_length:
        distance_cM = Gamma(k=gamma_k, theta=gamma_theta)
        distance_bp = distance_cM * 15000 * 100  # Convert cM to
bp
        position = position + distance_bp
        IF position < chromosome_length:
            crossover_positions.append(position)

    RETURN Sort(crossover_positions)
```

With default parameters (k=2, theta=0.38 cM), the mean inter-crossover distance is 0.76 cM, approximately 1.14 Mb.

#### B.4 Chromatid Generation

Given two parental haplotypes and crossover positions, four recombinant chromatids are generated. The model assumes no chromatid interference (any one of the two maternal chromatids can pair with any one of the two paternal chromatids at each crossover).

```
FUNCTION GenerateChromatids(parent1_haplotype, parent2_haplotype,
crossover_positions):

    # Create four chromatids: two copies of each parental
haplotype
    chromatids = [
        Copy(parent1_haplotype),  # chromatid 0
        Copy(parent1_haplotype),  # chromatid 1
        Copy(parent2_haplotype),  # chromatid 2
        Copy(parent2_haplotype)   # chromatid 3
    ]

    FOR each crossover_position IN crossover_positions:
        # Randomly select two chromatids to exchange (one from
each homolog)
        chromatid_A = RandomChoice([0, 1])      # From parent 1
        chromatid_B = RandomChoice([2, 3])      # From parent 2

        # Exchange genetic material distal to crossover point
        SwapDistalSegments(chromatids[chromatid_A],
                           chromatids[chromatid_B],
                           crossover_position)
```

```
    RETURN chromatids
```

#### B.5 Progeny Assembly

Each oocyst produces four genetically distinct haploid progeny through independent assortment of chromatids across chromosomes.

```
FUNCTION AssembleProgeny(parent1_genome, parent2_genome, gamma_k,
gamma_theta):

    progeny_genomes = [EmptyGenome() for i in 1 to 4]

    FOR each chromosome IN 1 to 14:

        # Get parental haplotypes for this chromosome
        p1_haplotype = parent1_genome.GetChromosome(chromosome)
        p2_haplotype = parent2_genome.GetChromosome(chromosome)

        # Place crossovers and generate chromatids
        crossovers = PlaceCrossovers(chromosome.length, gamma_k,
gamma_theta)
        chromatids = GenerateChromatids(p1_haplotype,
p2_haplotype, crossovers)

        # Randomly assign chromatids to progeny (without
replacement)
        shuffled_indices = RandomPermutation([0, 1, 2, 3])
        FOR i = 0 TO 3:
            progeny_genomes[i].SetChromosome(chromosome,
chromatids[shuffled_indices[i]])

    RETURN progeny_genomes
```

#### B.6 Scaling to Sporozoites

Each oocyst produces thousands of sporozoites, but only four distinct genotypes exist among siblings from a single oocyst. The model stores sporozoites in cohorts organized by genotype:

```
FUNCTION GenerateSporozoites(progeny_genomes, num_sporozoites):

    # Each oocyst produces 4 distinct genotypes in equal
proportions
    sporozoite_cohorts = []
    sporozoites_per_genotype = num_sporozoites / 4

    FOR i = 0 TO 3:
        cohort = CreateCohort(progeny_genomes[i],
sporozoites_per_genotype)
        sporozoite_cohorts.append(cohort)


    RETURN sporozoite_cohorts
```

With default parameters (mean 4,000 sporozoites per oocyst), each of the four sibling genotypes is represented by approximately 1,000 sporozoites.

When a mosquito bites a human, sporozoites are sampled from the available cohorts using multinomial sampling proportional to cohort populations. This maintains the genetic composition of the sporozoite pool while allowing efficient simulation of transmission events.

#### B.7 Complete Meiosis Algorithm

```
FUNCTION PerformMeiosis(gametocytes, num_oocysts, gamma_k,
gamma_theta, spz_per_oocyst_dist):

    all_sporozoite_cohorts = []

    # Pair gametocytes to form zygotes
    pairs = PairGametocytes(gametocytes, num_oocysts)

    FOR each (male_genome, female_genome) IN pairs:

        # Generate four progeny genomes through recombination
```

```
        progeny = AssembleProgeny(male_genome, female_genome,
gamma_k, gamma_theta)

        # Determine sporozoite count for this oocyst
        num_spz = Sample(spz_per_oocyst_dist)

        # Generate sporozoite cohorts (4 cohorts per oocyst)
        cohorts = GenerateSporozoites(progeny, num_spz)
        all_sporozoite_cohorts.extend(cohorts)

    RETURN all_sporozoite_cohorts
```

### Appendix C: Model Calibration

#### C.1 Overview

We jointly calibrated 12 EMOD parameters governing within-host parasite dynamics, transmission, and acquired immunity against age-stratified clinical incidence and parasite prevalence data from three reference sites spanning a wide transmission gradient: Namawala, Tanzania (annual EIR 329); Dielmo, Senegal (201); and Ndiop, Senegal (20) [9,10]. Calibration followed a two-stage iterative history-matching approach using machine-learning emulators [11,12]. Candidate parameter sets were first filtered for plausibility against site-level annual entomological inoculation rate (EIR), then EIR-plausible candidates were evaluated against age-stratified prevalence and clinical incidence likelihoods.

Calibration simulations used populations of 1,000 individuals with vital dynamics configured to maintain a stable sub-Saharan African age distribution. Each calibration simulation was run for 60 years, with outcome metrics averaged over the final three years. These calibration simulations were used to select epidemiological parameter values for the study scenarios; parasite genetic metrics were not used as calibration targets.

#### C.2 Calibrated parameters

The calibrated parameters and their prior bounds are listed in Table 2. Seven parameters control within-host parasite dynamics and transmission, and five control acquired immunity. Bounds were set to span physiologically plausible ranges informed by EMOD documentation and prior calibration experience [13,14]. Log-uniform priors were used for parameters spanning multiple orders of magnitude.

S1 Appendix Table 2: EMOD calibration parameters and priors

| Parameter | EMOD definition | Bounds | Prior | Final value |
|---|---|---|---|---|
| Antigen_Switch_Rate | Antigenic switching rate per infected red blood cell per asexual cycle | [1e-11, 1e-8] | Log-uniform | 1.56e-10 |
| Falciparum_PfEMP1_Variants | Number of distinct PfEMP1 variants in the parasite population | [600, 20000] | Uniform | 6932 |
| Base_Gametocyte_Production_Rate | Fraction of infected red blood cells producing gametocytes | [0.01, 0.2] | Log-uniform | 0.171 |
| Base_Gametocyte_Mosquito_Survival_Rate | Baseline fraction of gametocytes in a blood meal that successfully infect the mosquito | [1e-4, 0.9] | Log-uniform | 0.0350 |
| Fever_IRBC_Kill_Rate | Maximum rate at which fever clears infected red blood cells | [0.01, 1000] | Log-uniform | 1.19 |
| Antibody_Days_To_Long_Term_Decay | Days since last stimulation before antibody shifts to long-term decay | [365, 10000] | Log-uniform | 3365 |

| | | | | |
|---|---|---|---|---|
| Antibody_Long_Term_ Decay_Days | Exponential decay timescale of antibody titer under long-term decay | [365, 10000] | Log-uniform | 3365 |
| Antibody_IRBC_Kill_Rate | Scale factor converting antibody level to IRBC clearance rate | [0.5, 512] | Log-uniform | 1582 |
| Nonspecific_Antigenicity_ Factor | Weight on antibody-mediated clearance of antigenically weak surface proteins | [0.01, 1.0] | Log-uniform | 0.0939 |
| Nonspecific_Antibody_ Growth_Rate_Factor | Weight on antibody capacity growth for minor epitopes | [0.05, 100] | Log-uniform | 0.481 |
| Max_MSP1_ Antibody_Growthrate | Maximum increase in MSP1 antibody capacity per asexual cycle | [0.005, 1.0] | Log-uniform | 0.00516 |
| Falciparum_MSP_Variants | Number of distinct MSP variants in the parasite population | [10, 1000] | Log-uniform (rounded to integer) | 493 |

#### Seasonal EIR forcing

Monthly EIR patterns at each reference site were specified using linear-spline habitat multipliers adjusted through an autoregressive transfer function. EMOD's larval dynamics introduce temporal lag and smoothing between habitat forcing and realized EIR. We approximated this relationship using a seventh-order autoregressive model, in which realized monthly EIR depends on habitat forcing in the current and preceding six months. We then inverted this relationship to identify habitat forcing profiles that reproduced each site's target monthly EIR pattern. The AR kernel achieved monthly EIR shape correlations ≥0.80 across sites, indicating that it captured the dominant model-intrinsic lag between habitat availability and realized transmission.

#### EIR filtering

We used an iterative generate-filter-simulate approach over 14 rounds of approximately 700 simulations each, for approximately 9,800 simulations total. Two emulators were used in a consensus filtering arrangement: XGBoost [15] for fast coarse filtering and a sparse variational Gaussian process [16] trained on near-target simulations to provide probabilistic acceptance. Candidate parameter sets were retained if they produced annual EIR within approximately 20% of the reference values at all three sites simultaneously.

#### Likelihood evaluation

EIR-plausible candidates were evaluated against age-stratified prevalence and clinical incidence data. Prevalence was evaluated using a beta-binomial likelihood, and incidence was evaluated using a Bayesian posterior predictive negative-binomial likelihood following McCarthy et al (2015) [13]. Candidates were ranked using a weighted composite likelihood across the three reference sites, with threefold weight assigned to the Namawala prevalence likelihood. The top-ranked parameter set was used for the scenario simulations analyzed in the main text.

S1 Appendix Figure 4: Plausible EIR candidates compared to prevalence and clinical incidence.

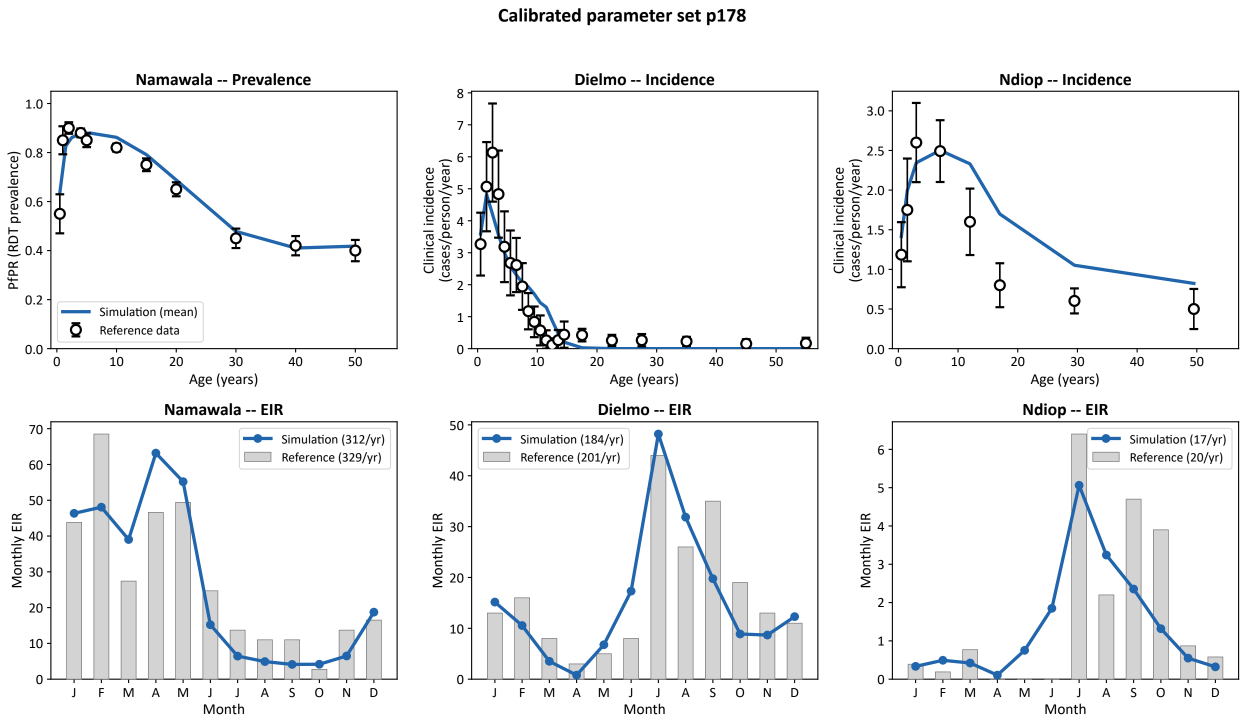

### Appendix D: Observational sampling model and subpopulation grouping

#### D.1 Population stratification options

The observational sampling model selects infections from EMOD infection-level outputs to mimic field parasite genetic data collection. Sampling eligibility can be defined using any individual- or infection-level variables recorded in the EMOD infection report, including time, node, age, clinical status, and infection complexity. Summary statistics can be calculated for the full sampled population or for user-defined subpopulations.

Monthly metrics are calculated from eligible infections within each monthly time window. To avoid repeated representation of persistent infections, an individual can contribute at most one sampled infection record per month. Yearly metrics are calculated from eligible monthly infection records after applying the specified annual sampling scheme. Pairwise relatedness metrics are calculated at the yearly scale by default for computational tractability.

S1 Appendix Table 3: Observational sampling options[1].

| Sampling Scheme | Description | Parameters | Status |
|---|---|---|---|
| **Random sampling** | Samples $N_{infections}$ per year with distribution mirroring true incidence trends across the year | $N_{Infections}$ per year | Base option |
| Multi-population proportions | Proportion of samples per population for even sampling or bias toward source/sink populations | Proportion per population, will split $N_{infections}$ per year proportionally | Optional |
| Monogenomic bias | Proportion of samples that are monogenomic vs. polygenomic to replicate known sequencing bias; can also be set as a subpopulation comparison | Proportion monogenomic/polygenomic for $N_{infections}$ per year | Optional |
| **Temporal sampling – seasonal** [2] | Samples $N_{infections}$ during wet or dry seasons up to available infections to match operational activities. | Seasonal length (full season or peak season) | Optional |
| **Temporal sampling – equal monthly** | Samples equal number of infections per month | | Optional |

[1] For the total number of infections per year or proportional split the calculations will provide the expected number of infections but may return fewer infections per group if fewer infections exist per group.

[2] Auto seasonality assignment from Sahelian patterns; adaptable to other time groupings.

#### D.2 Annual sampling schemes

For yearly analyses, the sampling model draws $N_{\text{infections}}$ infection records from the eligible monthly pool. When sampling uniformly from all eligible infections across the year, the resulting sample distribution follows the simulated temporal distribution of infections. If fewer eligible infections are available than requested for a sampling stratum, all available infections are returned, and the realized sample size is smaller than $N_{\text{infections}}$.

#### D.3 Annual sampling schemes

Temporal sampling windows can be defined using calendar months or relative to the seasonal incidence peak. In the analyses presented here, peak-season sampling used a three-month window centered on the month of highest clinical incidence, full-transmission-season sampling used a wider window spanning two months before through two months after the peak month, and full-year sampling included all eligible infections in a 12-month period.

#### D.4 Subpopulation comparison options

Subpopulation comparisons are defined by columns in the EMOD infection report. The same genetic metrics can be calculated separately within each subpopulation to assess how sampling from demographic or clinical groups changes estimated parasite genetic summaries.

S1 Appendix Table 4: Observational subpopulation comparison options.

| Comparison Type | Demographic Variable | Description | Use Case | Parameters | Status |
|---|---|---|---|---|---|
| **Sampling variability** | N/A | Metrics calculated across user-specified number of samplings to assess variability of $N_{infections}$ within population | Quantify sampling uncertainty | Number of sampling replicates | Optional (requires >1 sampling) |
| **Age distribution** | Age groups as a proxy of age-dependent immunity | Genetic metrics compared across age bins (default: 0-5, 5-15, 15+ years) | Evaluate generalizability of targeted populations (e.g., school-based surveys) to broader population | Age bin bounds and labels (user-configurable) | Optional |
| **Clinical presentation** | Symptomatic status as a proxy for care seeking | Metrics compared between individuals with and without fever | Assess clinical bias in sampling | Fever presence/absence | Optional |
| **Infection complexity** | COI (monogenomic vs. polygenomic) | Genetic metric trends compared between monogenomic and polygenomic infections | Evaluate sequencing strategy trade-offs: information content from monogenomic samples vs. polygenomic sensitivity improvements | COI classification | Optional |

#### References


1. Eckhoff PA. A malaria transmission-directed model of mosquito life cycle and ecology. Malar J. 2011;10: 303. doi:10.1186/1475-2875-10-303

2. Wong W, Wenger EA, Hartl DL, Wirth DF. Modeling the genetic relatedness of *Plasmodium falciparum* parasites following meiotic recombination and cotransmission. PLoS Comput Biol. 2018;14: e1005923. doi:10.1371/JOURNAL.PCBI.1005923

3. Miles A, Iqbal Z, Vauterin P, Pearson R, Campino S, Theron M, et al. Indels, structural variation, and recombination drive genomic diversity in *Plasmodium falciparum*. Genome Res. 2016;26: 1288–1299. doi:10.1101/GR.203711.115/-/DC1

4. Aurrecoechea C, Brestelli J, Brunk BP, Dommer J, Fischer S, Gajria B, et al. PlasmoDB: A functional genomic database for malaria parasites. Nucleic Acids Res. 2009;37. doi:10.1093/nar/gkn814

5. Bradley J, Stone W, Da DF, Morlais I, Dicko A, Cohuet A, et al. Predicting the likelihood and intensity of mosquito infection from sex specific *Plasmodium falciparum* gametocyte density. Elife. 2018;7. doi:10.7554/ELIFE.34463

6. Beier JC, Davis JR, Vaughan JA, Noden BH, Beier MS, Beier JC, et al. Quantitation of *Plasmodium falciparum* sporozoites transmitted in vitro by experimentally infected *Anopheles gambiae* and *Anopheles stephensi*. Am J Trop Med Hyg. 1991;44: 564–570. doi:10.4269/AJTMH.1991.44.564

7. Rosenberg R, Rungsiwongse J, Rosenberg R, Rungsiwongse J. The number of sporozoites produced by individual malaria oocysts. Am J Trop Med Hyg. 1991;45: 574–577. doi:10.4269/AJTMH.1991.45.574

8. Graumans W, Jacobs E, Bousema T, Sinnis P. When is a *Plasmodium*-infected mosquito an infectious mosquito? Trends Parasitol. 2020;36: 705–716. doi:10.1016/J.PT.2020.05.011

9. Smith TA, Charlwood JD, Kihonda J, Mwankusye S, Billingsley P, Meuwissen J, et al. Absence of seasonal variation in malaria parasitaemia in an area of intense seasonal transmission. Acta Trop. 1993;54: 55–72. Available: http://linkinghub.elsevier.com/retrieve/pii/0001706X9390068M

10. Rogier C, Tall A, Diagne N, Fontenille D, Spiegel A, Trape JF. *Plasmodium falciparum* clinical malaria: lessons from longitudinal studies in Senegal. Parassitologia.

1999;41: 255–259. Available: https://horizon.documentation.ird.fr/exl-doc/pleins_textes/pleins_textes_7/b_fdi_53-54/010020970.pdf

11. Vernon I, Gosling JP. A Bayesian computer model analysis of robust Bayesian analyses. Bayesian Anal. 2023;18: 1367–1399. doi:10.1214/22-BA1340

12. Andrianakis I, Vernon IR, McCreesh N, McKinley TJ, Oakley JE, Nsubuga RN, et al. Bayesian history matching of complex infectious disease models using emulation: A tutorial and a case study on HIV in Uganda. PLoS Comput Biol. 2015;11. doi:10.1371/journal.pcbi.1003968

13. McCarthy KA, Wenger EA, Huynh GH, Eckhoff PA. Calibration of an intrahost malaria model and parameter ensemble evaluation of a pre-erythrocytic vaccine. Malar J. 2015;14: 6. Available: http://eutils.ncbi.nlm.nih.gov/entrez/eutils/elink.fcgi?dbfrom=pubmed&id=25563798&retmode=ref&cmd=prlinks

14. Selvaraj P, Wenger EA, Gerardin J. Seasonality and heterogeneity of malaria transmission determine success of interventions in high-endemic settings: a modeling study. BMC Infect Dis. 2018;18: 413. doi:10.1186/s12879-018-3319-y

15. Chen T, Guestrin C. XGBoost: A scalable tree boosting system. Proceedings of the ACM SIGKDD International Conference on Knowledge Discovery and Data Mining. 2016;13-17-August-2016: 785–794. doi:10.1145/2939672.2939785;CSUBTYPE:STRING:CONFERENCE

16. Gardner JR, Pleiss G, Bindel D, Weinberger KQ, Wilson AG. GPyTorch: Blackbox matrix-matrix Gaussian process inference with GPU acceleration. Adv Neural Inf Process Syst. 2018;31. Available: https://gpytorch.ai.
